# Combined radiomics and liquid biopsy reflect tumor biology towards multimodal non-invasive prostate cancer risk stratification

**DOI:** 10.1101/2025.09.06.25335220

**Authors:** Anja Lisa Riediger, Clara Meinzer, Daniela Janscho, Samaneh Eickelschulte, Florian Janke, Olga Lazareva, Daniel Hübschmann, Oliver Stegle, Stefan Fröhling, David Bonekamp, Heinz-Peter Schlemmer, Holger Sültmann, Magdalena Görtz

## Abstract

Accurate risk stratification is essential to prevent over- and undertreatment in newly diagnosed prostate cancer (PCa). In a pilot study with 49 men (36 PCa patients, 13 controls), we integrated non-invasive data modalities collected in a real-world clinical setting: radiomics from multiparametric magnetic resonance imaging (T2/ADC index-lesion features), routine and extended serological parameters, multimodal liquid biopsy features (copy number variations, chromosomal instability, methylation, fragmentation in plasma/urinary cfDNA), plus clinical and lifestyle factors. Pairwise Spearman analyses revealed significant inter-modality correlations. Our holistic PCa characterization showed that imaging phenotypes reflect tumor stage and molecular aggressiveness: lower ADC metrics correlated inversely with genomic cfDNA features, while ADC/T2 lesion volumes correlated positively with molecular signals. These results indicate that routine imaging and serological data may capture systemic tumor biology. Our proof-of-concept demonstrates feasibility of combining multiple data modalities in real-world clinical workflows and supports developing integrative models for non-invasive PCa risk stratification.

## Introduction

Prostate cancer (PCa) is a leading cancer diagnosis among men^1^ and represents a significant clinical and societal burden, owing to the difficulty in distinguishing aggressive from indolent forms^2,3^. The limitations of current screening and risk assessment tools contribute significantly to this diagnostic uncertainty^4^. Prostate-specific antigen (PSA) testing in the blood plays a pivotal role for PCa screening, but suffers from limited specificity, particularly within intermediate concentration ranges, which frequently leads to avoidable prostate biopsies^5–7^. This challenge contributes to both underdiagnosis and overtreatment, leading to variability in disease course and clinical outcomes^8,9^. Clinical guidelines recommend the use of multivariable risk calculators that incorporate PSA derivatives alongside other clinical factors, such as a positive family history^3,10^. These tools aim to improve diagnostic precision, in order to support personalized risk assessment, guide shared decision-making, and reduce unnecessary subsequent examinations by identifying men at low risk who may safely undergo clinical follow-up. However, they still lack sufficient accuracy to reliably distinguish clinically significant from indolent PCa^2,3^. Multiparametric magnetic resonance imaging (mpMRI) and targeted biopsy have further enhanced the accuracy of PCa detection^11,12^. Prostate biopsy remains the diagnostic gold standard, but it is inherently invasive, associated with patient discomfort and procedural risks, and may fail to adequately capture tumor heterogeneity^13^. Given these limitations, increasing attention has been directed toward novel noninvasive strategies aimed at enhancing diagnostic accuracy and risk stratification. Liquid biopsy (LBx) has emerged as a promising approach for minimally invasive tumor profiling and disease monitoring through the analysis of circulating DNA and other blood- or urine-derived biomarkers^14,15^. Furthermore, serological parameters related to systemic inflammation, metabolism, and hormonal status have been associated with PCa risk and aggressiveness^16–19^.

Recent studies also highlight the relevance of modifiable risk factors, such as diet and lifestyle, in the etiology and progression of PCa^20–22^. Finally, radiological features derived from the quantitative analysis of mpMRI provide additional, noninvasive insights into tumor characteristics and may improve diagnostic precision^23–25^. The growing field of radiogenomics integrates imaging metrics with genomic or transcriptomic signatures, providing surrogate imaging biomarkers for molecular phenotypes which enables non-invasive subtyping and potentially informs treatment response prediction in PCa^26,27^. Beyond tissue-based analyses, combining radiomics with circulating biomarkers offers a promising approach for non-invasive tumor characterization. For example, radiomic heterogeneity metrics derived from computed tomography predicted circulating tumor DNA (ctDNA) levels in metastatic melanoma, indicating that imaging and LBx may provide complementary information on tumor burden^28^. Integrating multiple noninvasive biomarkers into a unified diagnostic framework may offer a more comprehensive assessment of PCa characteristics, including indicators of tumor aggressiveness and progression risk. Applied within a routine clinical workflow, these approaches harbor potential to improve PCa risk stratification, support individualized decision-making, and reduce unnecessary invasive procedures. Furthermore, the accumulation of multi-source data offers an opportunity to develop “digital twins”, models that integrate clinical, imaging and molecular data to simulate patient-specific disease outcomes^29^.

In this pilot study, we demonstrate the feasibility of assembling and analyzing a real-world PCa screening dataset that is the first to integrate multiple non-invasive modalities within a single PCa cohort: LBx markers, routine and extended serological parameters, mpMRI-derived radiomics, and patient-reported lifestyle factors. We interrogate cross-modality correlations and assess the potential of this multimodal approach to strengthen future risk stratification in routine clinical practice. By integrating systemic, molecular, imaging-based, and behavioral dimensions of patient data, our framework provides proof of concept for advancing toward an individualized digital twin of PCa that enables multimodal characterization of disease biology.

## Results

### Patient characteristics

Forty-nine men underwent PCa screening, including PSA testing, mpMRI, and prostate biopsy to confirm or exclude PCa diagnosis (**Table 1**). PCa was diagnosed in 36 men with the majority (29 of 36; 80.6%) harboring localized PCa (lPCa; N0 M0), while seven advanced PCa (aPCa) patients (19.4%) presented with lymph node (N1) and/or distant metastases (M1). In 13 men, prostate biopsies revealed no histopathological evidence of malignancy, serving as cancer-free controls. Comprehensive clinical data from electronic health records were available for all participants. MpMRI served as imaging modality for identifying and characterizing suspicious prostate lesions, which were scored using the PI-RADS system^30^ to inform subsequent targeted and systematic prostate biopsies. Furthermore, first-order radiological features were assessed in defined index lesions.

**Table 1:** Overview of patients and available multimodal data.

| **PCa, localized disease (stage I-III) (n = 29)** |  |  |  |  |  |
| --- | --- | --- | --- | --- | --- |
| age, median (range) |  |  | 66 (49–79) |  |  |
| PSA level (ng/mL), median (range) |  |  | 7.9 (1.8–28.7) |  |  |
|  | <u>LBx data</u> |  | radiomics data | serolomics data | survey data |
|  | plasma | urine |  |  |  |
| <u>low risk</u> | <u>4</u> | <u>3</u> | <u>3</u> | <u>4</u> | <u>4</u> |
| GS 6 (n = 4) | 4 | 3 | 3 | 4 | 4 |
| <u>intermediate risk</u> | <u>19</u> | <u>17</u> | <u>18</u> | <u>18</u> | <u>19</u> |
| GS 7a (n = 17) | 17 | 15 | 16 | 16 | 17 |
| GS 7b (n = 2) | 2 | 2 | 2 | 2 | 2 |
| <u>high-risk</u> | <u>6</u> | <u>6</u> | <u>6</u> | <u>6</u> | <u>5</u> |
| GS 7b (n = 1) | 1 | 1 | 1 | 1 | 0 |
| GS 8 (n = 1) | 1 | 1 | 1 | 1 | 1 |
| GS 9 (n = 4) | 4 | 4 | 4 | 4 | 4 |
| **PCa, disseminated disease (stage IV) (n = 7)** |  |  |  |  |  |
| age, median (range) |  |  | 66 (58–75) |  |  |
| PSA level (ng/mL), median (range) |  |  | 29.7 (6.3–391.3) |  |  |
|  | <u>LBx data</u> |  | radiomics data | serolomics data | survey data |
|  | plasma | urine |  |  |  |
| <u>lymph node metastases, N1 M0</u> | <u>3</u> | <u>3</u> | <u>2</u> | <u>3</u> | <u>2</u> |
| GS 7a (n = 1) | 1 | 1 | 1 | 1 | 0 |
| GS 8 (n = 1) | 1 | 1 | 1 | 1 | 1 |
| GS 9 (n = 1) | 1 | 1 | 0 | 1 | 1 |
| <u>distant metastases, M1</u> | <u>4</u> | <u>4</u> | <u>4</u> | <u>4</u> | <u>4</u> |
| GS 7a (n = 1) | 1 | 1 | 1 | 1 | 1 |
| GS 8 (n = 2) | 2 | 2 | 2 | 2 | 2 |
| GS 9b (n = 1) | 1 | 1 | 1 | 1 | 1 |
| **cancer-free participants (n = 13)** |  |  |  |  |  |
| age, median (range) |  |  | 60 (57–74) |  |  |
| PSA level (ng/mL), median (range) |  |  | 5.6 (4.2–14.4) |  |  |
|  | <u>LBx data</u> |  | radiomics data | serolomics data | survey data |
|  | plasma | urine |  |  |  |
| <u>controls with no evidence of malignancy in prostate biopsy</u> | 13 | 12 | 9 | 13 | 12 |

PCa stages were determined based on UICC (stage I-IV)<sup>32</sup> and D'Amico Risk Classification<sup>33</sup>
Abbreviations: LBx = Liquid Biopsy, M0/M1 = absence/presence of distant metastases, MRI = magnetic resonance imaging, N1 = presence of lymph node metastases, GS = Gleason Score, NA = data not available, n = number, PCa = prostate cancer, PSA = prostate-specific antigen, UICC = Union for International Cancer control

Blood samples were collected for both routine laboratory testing and extended serological analyses. Additional paired blood and urine samples were obtained for comprehensive LBx analyses (**Table 1**). Results from genomic and epigenomic cell-free DNA (cfDNA) profiling were available from previously published work^31^, in which a larger LBx cohort was analyzed. The present analyses included the subset of participants overlapping with the current multimodal dataset. LBx analyses included genome-wide methylation and copy number variation (CNV) profiling, chromosomal instability analysis (CIA), and the assessment of cfDNA fragmentation. For each participant, eight LBx features were assessed: estimated tumor fraction (TFx) based on genome-wide CNVs, CIA score, methylation score in PCa-specific regions, as well as two cfDNA fragmentation metrics (10 bp-oscillation score in plasma, and the proportion of cfDNA fragments with 163-169 bp lengths in urine (P163–169 bp)).

During sample collection, all participants received a structured survey covering lifestyle characteristics (e.g., meat and alcohol consumption, smoking history, physical activity), family history of PCa (first- and second-degree relatives), and known or suspected PCa risk factors, such as history of infertility or sterilization, occurrence of urinary tract infection or urinary retention within the previous six months (*Supplementary Table S1*). Complete survey data were available for all 13 cancer-free participants and 27 PCa patients; in 9 PCa cases, individual responses were missing. No significant differences in lifestyle or risk factors were observed between PCa patients and cancer-free participants (*Supplementary Table S1*). Similarly, age did not significantly differ between the two groups.

Overall, four diagnostic modalities (LBx, electronic health records, serolomics, radiomics) were collected within the clinical workflow (**Figure 1**), and complete annotated datasets were available for 42 men (33 PCa patients and 9 cancer-free participants).

**Figure 1:**
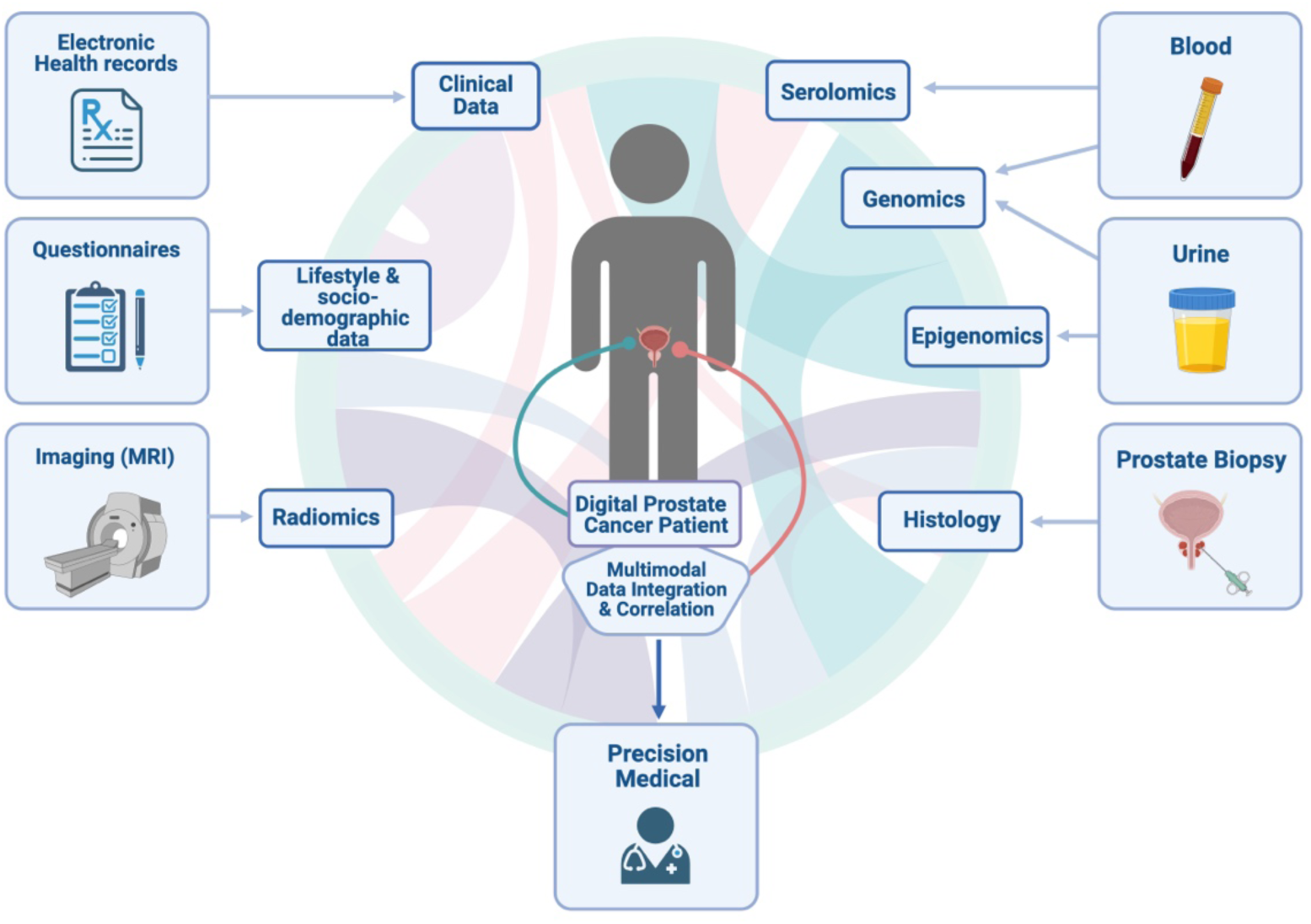
Multimodal data collection in a real-world clinical setting. Overview of multimodal data sources collected within the clinical workflow to achieve a holistic characterization of PCa patients. Clinical data (from electronic health records), lifestyle and socio-demographic data (from questionnaires), and radiomics features (from mpMRI) were correlated with molecular and pathological modalities, including serolomics (from blood), genomic and epigenomic cfDNA profiling (from blood and urine), and histological information (from prostate biopsy). As a future perspective, integration of these multimodal datasets has the potential to enable the development of digital PCa patient profiles (“digital twins”) to support clinical decision making and precision medicine approaches. Abbreviations: cfDNA = cell-free DNA, mpMRI = multiparametric magnetic resonance imaging, PCa = prostate cancer

### Radiological features of prostate lesions distinguished between PCa patients and cancer-free individuals

Annotated prostate MRI data were available for 44 of the 49 men, including 35 PCa patients and 9 cancer-free controls (**Table 1**). Radiological data were incomplete or missing for one patient with advanced PCa (N1M0) and four cancer-free controls. Suspicious prostate lesions visible on mpMRI were annotated by experienced radiologists according to PI-RADS criteria^30^. These lesions were subsequently targeted during image-guided biopsy, alongside standard systematic sampling. A total of 80 suspicious lesions were identified across the 44 men (median: 2 lesions per patient; range: 1–5). For each participant, one index lesion was defined for downstream analysis, and quantitative, first-order radiological features were obtained from T2-weighted imaging (T2) or apparent diffusion coefficient (ADC) mapping based on diffusion-weighted imaging. These features included intensity-based and volumetric measures such as mean, median, minimum, maximum, 10^th^-percentile, 90^th^-percentile, standard deviation, variance, and lesion volume. The index lesions underwent histopathological evaluation following targeted biopsy, providing the matched diagnostic reference standard for subsequent analyses. The radiological assessment of the selected index lesions demonstrated a sensitivity and specificity of 91%, respectively, in distinguishing malignant from benign lesions, as confirmed by histopathology (**Table 2**).

**Table 2:** Radiological and histopathological assessment of prostate index lesions.

|  |  | Histopathological tumor status |  |
| --- | --- | --- | --- |
| | | Malignant (n = 33):<br>Gleason ≥ 6 | Benign (n = 11):<br>no sign of malignancy |
| Predicted tumor status<br>(radiological<br>assessment) | Cancer likely:<br>PI-RADS 4 + 5 | 30 | 1 |
|  | Equivocal:<br>PI-RADS 3 | 3 | 10 |
|  |  | True positive: 30<br>False negative: 3 | True negative: 10<br>False positive: 1 |

Confusion matrix displaying the predicted tumor status by the radiological assessment based on the PI-RADS classification (cancer (highly) likely: PI-RADS 4 + 5; equivocal: PI-RADS 3) and the histopathological tumor status defined by the pathological evaluation based on the Gleason score (malignant: Gleason ≥ 6; benign: no pathological sign of malignancy)

Two PCa patients presented with PI-RADS 3 index lesions, which were confirmed as benign on targeted biopsy, and PCa diagnosis was identified based on systematic biopsies. As a result, the corresponding radiological features derived from the index lesions were considered non-informative for tumor characterization, and both cases were excluded from further radiological analyses.

The majority of the remaining 42 index lesions from both PCa patients and cancer-free individuals were located in the peripheral zone (PZ) of the prostate (n = 31), with four lesions showing additional invasion into the transition zone (TZ). Ten lesions were exclusively located in the TZ, and one TZ lesion showed an invasion into PZ. Statistical comparisons of radiological features extracted from prostate index lesions in PCa patients and cancer-free individuals revealed significant differences in several parameters (*Supplementary Table S2*), including T2 volume (adjusted p = 0.009), ADC volume (adjusted p = 0.023), ADC minimum (adjusted p = 0.034), as well as ADC 10^th^-percentile, mean, and median values (all adjusted p = 0.036; **Figure 2, a**). These features exhibited gradual trends across disease stages, with values either increasing (ADC volume, T2 volume) or decreasing (ADC minimum, 10^th^-percentile, median and mean) from cancer-free individuals to lPCa and further to aPCa patients (**Figure 2, b**).

**Figure 2:**
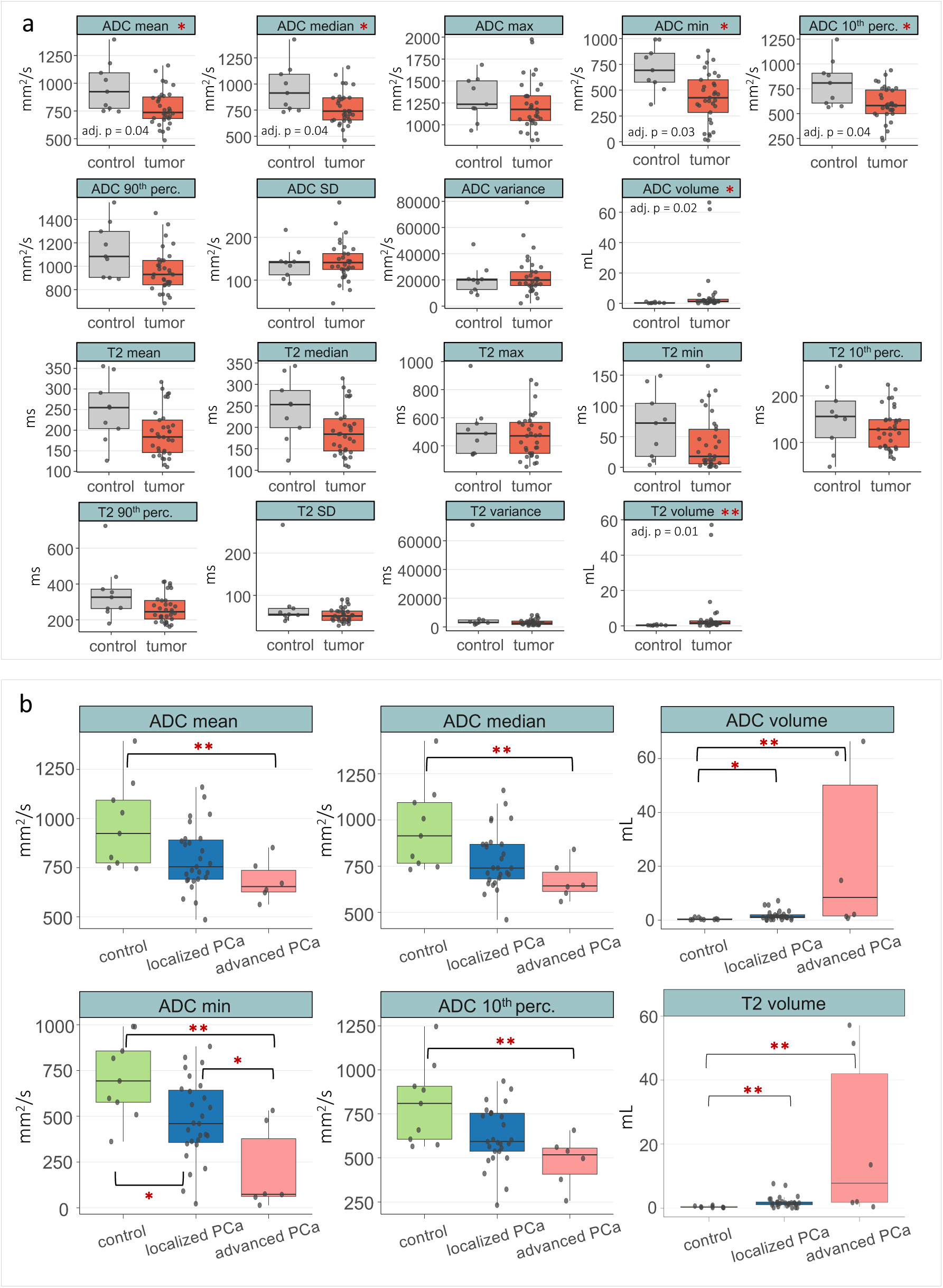
Assessment of first-order radiological features from mpMRI based on T2-weighted imaging or ADC mapping for 42 index lesions of 23 PCa patients and 9 cancer-free individuals. **a)** Comparison between all PCa patients and cancer-free individuals for all radiological features. **b)** Comparison between cancer-free individuals, and tumor patients stratified for lPCa and aPCa for all radiological features with significant differences between all PCa patients and cancer-free individuals. **a + b)** Box plot center lines indicate the median, and boxes illustrate the interquartile range with Tukey whiskers. Each dot represents one sample. Significant differences (adjusted p < 0.05) are highlighted with an asterisk (* p <0.05, ** p <0.01). Abbreviations: ADC = apparent diffusion coefficient, aPCa = advanced PCa, lPCa = localized PCa, max = maximum, min = minimum, perc. = percentile, SD = standard deviation

Statistical comparisons between the three subgroups revealed significantly lower values for ADC 10^th^-percentile, mean, median, and minimum in aPCa patients compared to cancer-free individuals (adjusted p = 0.002–0.009; **Figure 2, b**; *Supplementary Table S3*). The stepwise decline of ADC minimum values was further reflected by significant differences between aPCa and lPCa patients (adjusted p = 0.043), as well as between lPCa patients and cancer-free individuals (adjusted p = 0.040). Similar trends were also observed for T2-derived metrics (*Supplementary Table S3 and Supplementary Figure S1*). Advanced PCa patients showed significantly reduced T2 mean, median, and 90^th^-percentile values compared to cancer-free individuals (adjusted p = 0.035 – 0.042). Additionally, a stepwise decrease was observed for T2 minimum values across disease stages, with aPCa patients harboring significantly lower values compared to both lPCa patients (adjusted p = 0.038) and cancer-free individuals (adjusted p = 0.018), while lPCa patients did not significantly differ from cancer-free individuals.

Conversely, T2- and ADC-derived tumor volumes increased progressively across disease stages, highlighting the progressive increase in lesion size. Both lPCa and aPCa patients demonstrated significantly larger lesion volumes compared to cancer-free individuals (T2 volume: adjusted p = 0.006 and 0.001, respectively; ADC volume: adjusted p = 0.021 and 0.002, respectively; **Figure 2, b**; *Supplementary Table S3*).

Overall, these radiological parameters differentiated PCa patients from cancer-free individuals and showed consistent trends across disease stages, indicating their association with tumor stage.

### Comprehensive laboratory profiling highlighted PSA-associated and hormonal changes in PCa patients

Extended routine laboratory profiling was performed for all 49 men, encompassing 31 blood-based parameters related to lipid metabolism, inflammation, hormonal regulation, and vitamin status. Two men with lPCa had missing laboratory values due to technical issues, resulting in complete datasets for 47 participants (**Table 1**).

To assess differences in serological markers, all PCa patients were initially compared to cancer-free individuals (*Supplementary Table S4 and Supplementary Figures S2–4, a*). Among all 31 evaluated parameters, PSA density emerged as the most robust discriminator, with significantly elevated levels in PCa patients (0.19 ng/mL^2^ vs. 0.09 ng/mL^2^; adjusted p = 0.027; **Figure 3, a**). Additional markers showing nominal significance (unadjusted p < 0.01; all adjusted p = 0.064) included the absolute monocyte count, androstenedione, DHEA-sulfate (DHEA-S), and the free-to-total PSA ratio (free PSA%) (**Figure 3, a**). PCa patients exhibited lower median levels of androstenedione (66.5 vs. 94.0 ng/dL) and DHEA-S (0.8 vs. 1.4 µg/mL), and free PSA% (15.6% vs. 24%), while absolute monocyte counts were elevated compared to cancer-free individuals (0.4 vs. 0.3 /nL).

**Figure 3:**
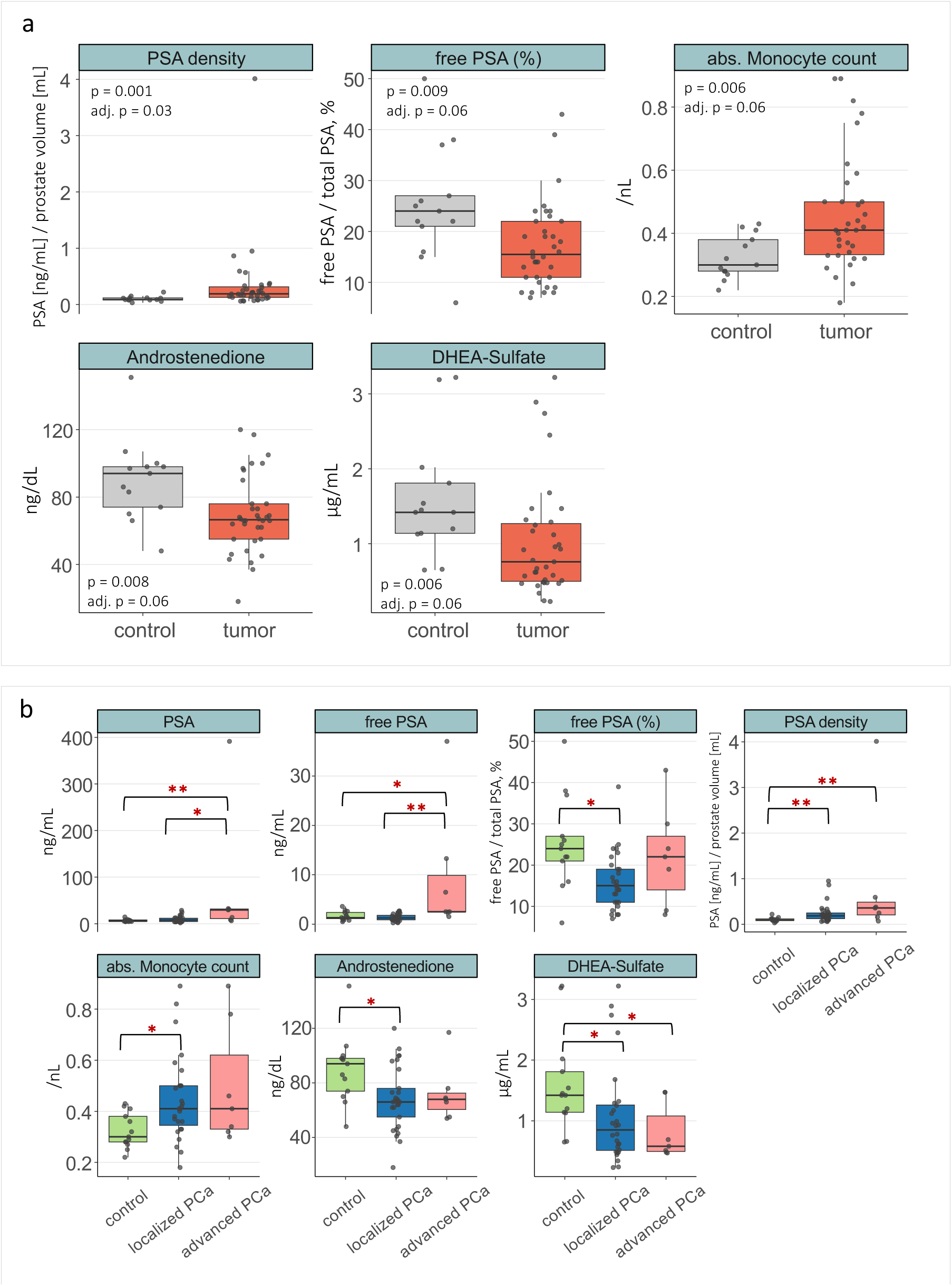
Assessment of 31 laboratory parameters measured in the blood from PCa patients and cancer-free individuals. **a)** Comparison between all PCa patients and cancer-free individuals for all laboratory parameters with notable differences between the two groups. **b)** Comparison between cancer-free individuals, and tumor patients stratified for lPCa and aPCa for all laboratory parameters with significant differences between the three cohorts. **a + b)** Box plot center lines indicate the median, and boxes illustrate the interquartile range with Tukey whiskers. Each dot represents one sample. Significant differences (adjusted p < 0.05) are highlighted with an asterisk (* p <0.05, ** p <0.01). Abbreviations: abs = absolute, adj. p = adjusted p value, DHEA-Sulfate = dehydroepiandrosterone sulfate, PSA = prostate specific antigen

To further explore stage-dependent trends, subgroup analyses were performed among cancer-free individuals, patients with lPCa, and those with aPCa (*Supplementary Table S5 and Supplementary Figures S2–4, b*). This extended analysis revealed that the PSA density increased steadily across disease stages, with medians of 0.09 ng/mL² in controls, 0.18 ng/mL² in lPCa, and 0.36 ng/mL² in aPCa (**Figure 3, b**). Both lPCa and aPCa patients showed significantly higher median values of the PSA density compared to controls (adjusted p = 0.008 and 0.004, respectively), while the difference between lPCa and aPCa did not reach significance (adjusted p = 0.164). Total PSA also followed an increasing trend and was significantly elevated in aPCa compared to controls (29.7 ng/mL vs. 5.6 ng/mL; adjusted p = 0.008) and to lPCa (29.7 ng/mL vs. 7.9 ng/mL; adjusted p = 0.012), although lPCa patients did not differ significantly from controls (7.9 ng/mL vs. 5.6 ng/mL; adjusted p = 0.47). Free PSA% showed a non-linear trend, with highest values in controls (median = 24%), a significant decrease in lPCa (median = 15%; adjusted p = 0.012 for control vs. lPCa), and a partial increase in aPCa (median = 22%).

Among hormonal parameters, both DHEA-S and androstenedione levels showed a decreasing trend from cancer-free individuals to lPCa to aPCa patients (**Figure 3, b**). DHEA-S was significantly reduced in both lPCa and aPCa versus controls (0.9 µg/mL and 0.6 µg/mL vs. 1.4 µg/mL, respectively; adjusted p = 0.031 for both comparisons). Androstenedione was significantly lower in lPCa versus controls (66 ng/dL vs. 94 ng/dL; adjusted p = 0.022), though the difference was not significant for aPCa. Furthermore, testosterone, dihydrotestosterone and progesterone equally exhibited consistent decreasing trends from cancer-free individuals to lPCa and aPCa patients, while estradiol and sex hormone-binding globulin (SHBG) showed an opposite, increasing trend. However, none of these differences were statistically significant (*Supplementary Table S5 and Supplementary Figure S3, b*).

For immune-related markers, the absolute monocyte count was elevated in PCa patients, with significantly higher levels in lPCa compared to controls (0.4 v. 0.3 /nL; adjusted p = 0.026; **Figure 3, b**). Although aPCa patients showed a similar trend, the difference did not reach statistical significance after correction for multiple testing (0.4 v. 0.3 /nL; adjusted p = 0.065). Median values were comparable between lPCa and aPCa patients (both 0.4 /nL). The relative proportion of monocytes also demonstrated an increasing trend across disease stages, though differences were not statistically significant. Additional white blood cell parameters commonly linked to inflammatory activity, including absolute leukocyte and neutrophil counts, respectively, showed similar upward trends from cancer-free individuals to lPCa and aPCa patients (*Supplementary Table S5 and Supplementary Figure S4, b*). However, no significant differences were observed between subgroups. The assessment of metabolic laboratory parameters and vitamins did not reveal any distinct patterns between PCa patients and cancer-free individuals (*Supplementary Table S5 and Supplementary Figure S5, b*).

Overall, PSA-related parameters demonstrated the most consistent and significant stage-associated changes, while certain hormonal and immune cell markers also revealed notable trends across disease groups.

### Combined genomic and epigenomic plasma and urinary cfDNA profiling improved ctDNA detection

Plasma and urine samples from PCa patients were subjected to genomic and epigenomic cfDNA analyses for tumor characterization. For each patient, eight cfDNA-based features were assessed to enable ctDNA detection, as previously described^31^. The genomic measures included the estimated TFx and the CIA score, both derived from genome-wide CNV and chromosomal instability profiles. Epigenomic features comprised a PCa-specific methylation score and two particular cfDNA fragmentation characteristics in plasma (10 bp-oscillation score) and in urine (proportion of fragments with 163-169 bp length). Genomic and epigenomic analyses revealed both consistent and complementary information between matched plasma and urine samples. In lPCa, ctDNA detection rates based on single-parameter analyses were generally low (**Figure 4, a**). The highest ctDNA detection was achieved with the CIA score in urine (17%), followed by both the methylation score and the plasma cfDNA fragmentation feature (each 14%). Other parameters yielded detection rates below 10%. Advanced PCa patients demonstrated higher ctDNA detection across all features (**Figure 4, a**), with the CIA score in urine showing the strongest performance (57%), followed by the methylation score in urine and the plasma fragmentation feature (both 43%).

**Figure 4:**
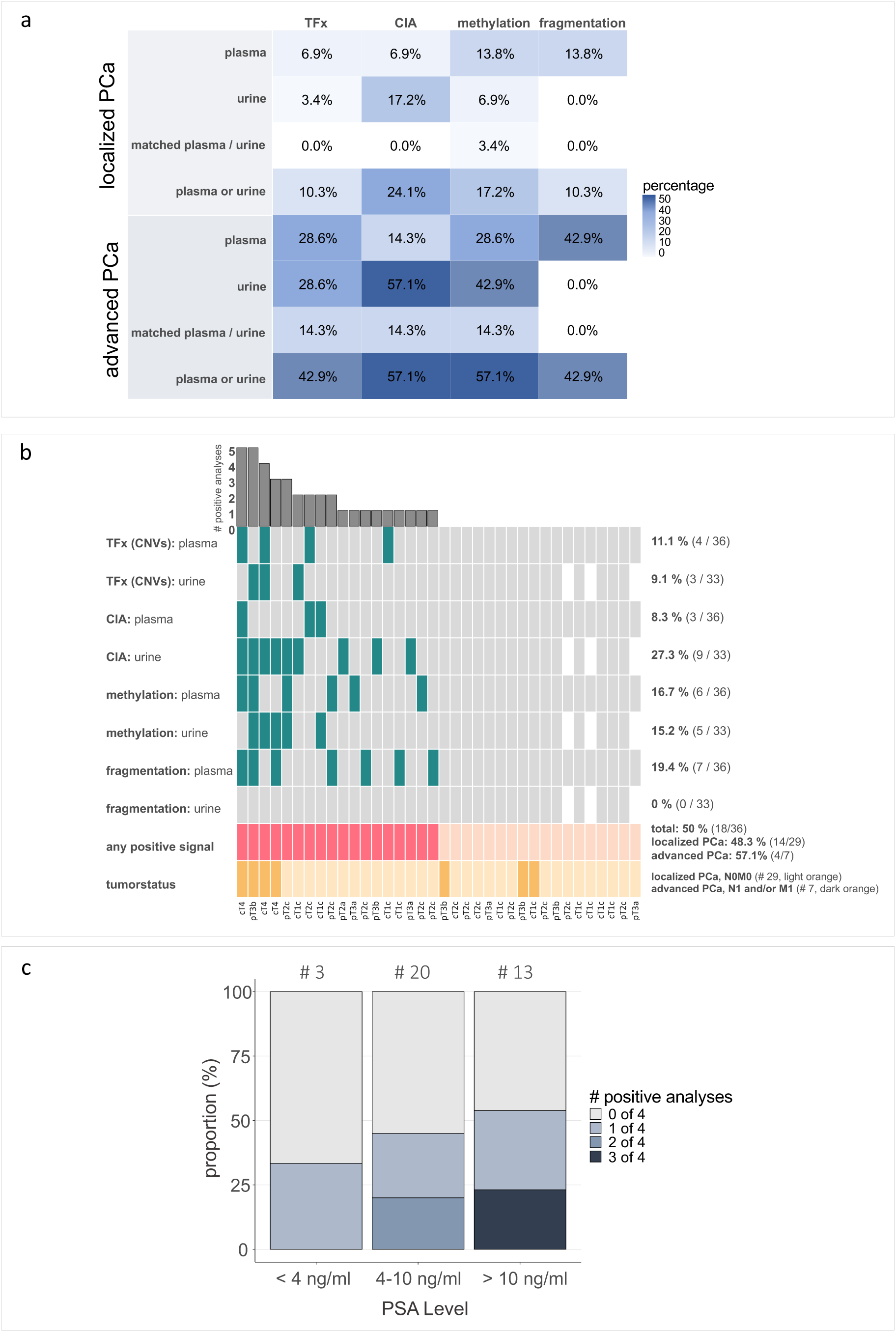
PCa detection and risk stratification based on multimodal LBx analyses. **a)** Detection rates of tumor signals (ctDNA) in plasma and urine samples from lPCa (top) and aPCa (bottom) patients based on four (epi)genomic analyses: 1) estimated TFx based on the CNV analysis 2) CIA score, 3) methylation score, 4) cfDNA fragmentation (10 bp-oscillation score in plasma cfDNA, P163–169 bp in urinary cfDNA). Detection rates (percentages, %) are shown separately for plasma and urine, in matched plasma and urine, and based on the complementary analysis combining results in both plasma and urine. Overall, 32 PCa patients harbored matched plasma and urine samples; 4 lPCa patients with missing urine samples. **b)** Oncoprint with results from genomic (estimated TFx based on CNVs, CIA score) and epigenomic (cfDNA fragmentation features, methylation score) analyses in plasma and urinary cfDNA. Patients with detectable ctDNA in the respective analyses are indicated with colored tiles. White tiles represent non-available urine samples (n = 4). The bar plot on top depicts the number of positive analyses per patient. The second-to-last row represents the number of PCa patients with positive signal in at least one analysis (“any positive signal”; colored tiles). The last row indicates the tumor status (lPCa and aPCa) and the tumor size according to the TNM classification. **c)** Association between the number of positive LBx analyses and the PSA levels for PCa patients. Distribution of patients based on the number of positive tumor signals detected (0–4 out of 4 analyses) in relation to PSA levels (<4 ng/mL, 4–10 ng/mL, and >10 ng/mL). Total numbers are depicted above each bar. Tumor signal positivity is determined using complementary analysis in plasma and urine across the four (epi)genomic assessments: TFx, CIA score, cfDNA fragmentation features, and methylation score. Abbreviations: CIA = chromosomal instability analysis, ctDNA = circulating tumor-derived DNA, M0/M1 = absence/presence of distant metastases, N0/N1 = absence/presence of lymph node metastases # = number of, TFx = tumor fraction, cT/pT = clinical/pathological T stage (tumor size), P163-169 bp = proportion of cfDNA fragments with 163– 169 bp lengths.

Combined analysis of matched plasma and urine improved ctDNA detection, particularly in lPCa (**Figure 4, a**). In this subgroup, the CIA score achieved the highest detection rate (24%) when both biofluids were considered. For aPCa, the highest detection rates were observed for both the methylation score and the CIA score (both 57%). Considering all eight genomic and epigenomic cfDNA features in plasma and urine, ctDNA was detectable in 50% of all PCa patients (18/36) based on at least one positive result (**Figure 4, b**). Subgroup analysis revealed ctDNA detection in 48% of lPCa and 57% of aPCa patients.

The ctDNA detection was not restricted to PCa patients with high PSA levels. CtDNA was also present in 45% of PCa patients with PSA levels 4–10 ng/mL and even in 33% of cases with PSA levels <4 ng/mL, based on at least one (epi)genomic cfDNA analysis type (**Figure 4, c**). However, the proportion of ctDNA-positive samples was increased among PCa patients with PSA levels exceeding 10 ng/ml. Notably, ctDNA was also identified in five PCa patients with UICC stage I tumors (T1c or T2a, all N0M0), one with Gleason 6, two with Gleason 7a, and one with Gleason 8 histology. In all five cases, PSA concentrations were below 10 ng/mL, with one patient showing levels below 4 ng/mL.

In comparison to the radiological assessment, PCa patients with a higher tumor burden and larger lesion volumes (**Figure 5, A**) tended to show stronger molecular signals in LBx samples than those with intermediate or small lesions (**Figure 5, B**). Consistently, PI-RADS 5 index lesions were associated with higher molecular tumor signals compared to PI-RADS 4 lesions. Among 18 PCa patients with detectable ctDNA in plasma and/or urinary cfDNA, 12 PCa patients presented with PI-RADS 5 index lesions, while four PCa patients harbored a PI-RADS 4 index lesion. One PCa patient with detectable ctDNA had no corresponding radiological data available. A positive molecular tumor signal based on the plasma cfDNA methylation analysis was also detected in one PCa patient with an equivocal PI-RADS 3 lesion (**Figure 5, C).** The anatomical location of the tumor also appeared to influence molecular signal distribution in blood and urine. Tumors with bladder invasion demonstrated high tumor signals in urine, whereas tumors located with larger distance to the urethra or bladder preferentially harbored signals in plasma, as illustrated by two representative cases (**Figure 5, A + D**).

**Figure 5:**
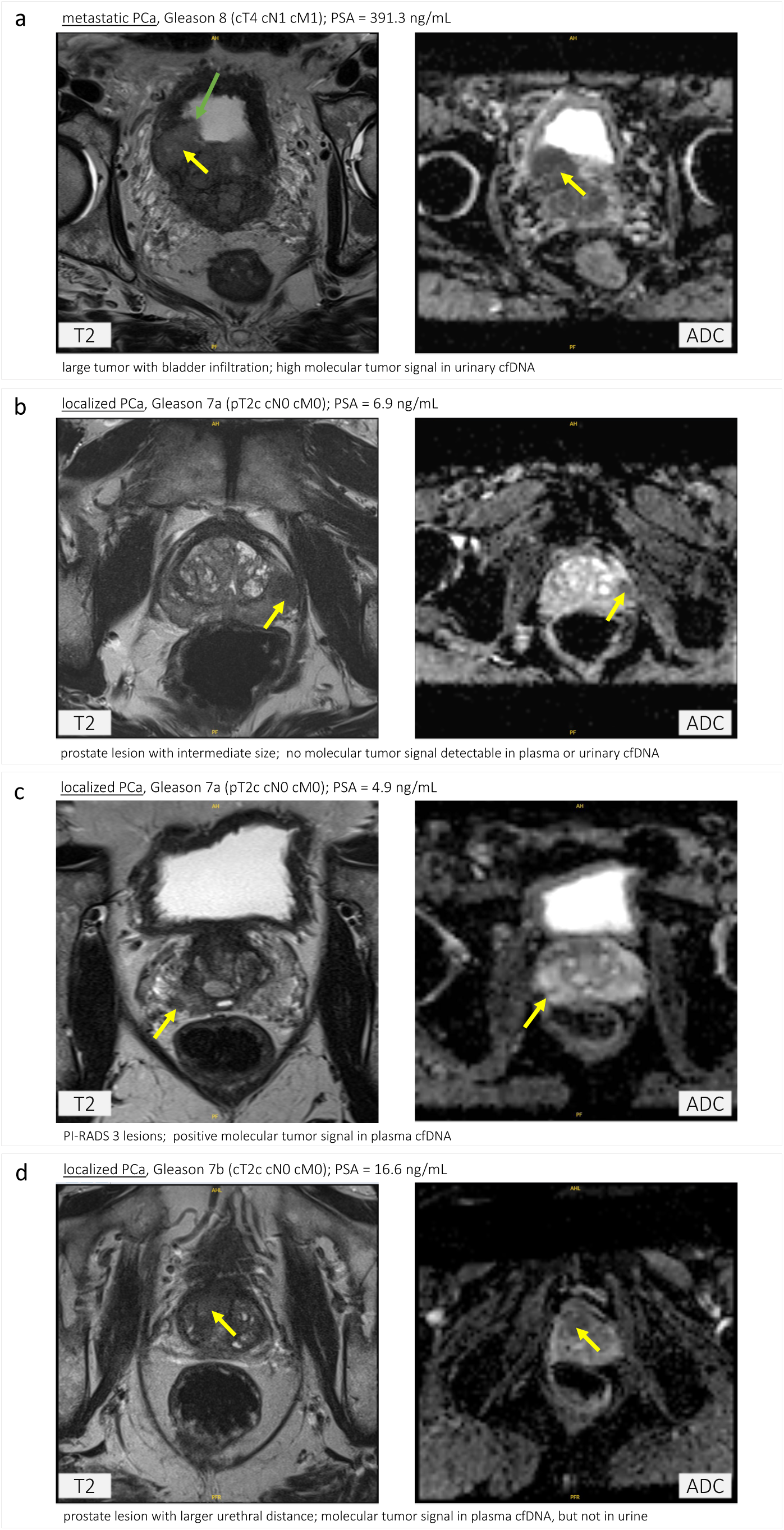
Radiological examples and corresponding LBx signals in PCa patients. Representative mpMRI images (T2-weighted sequence, left; ADC mapping, right) from four PCa patients demonstrating the association between radiological tumor characteristics and molecular tumor signals in plasma and/or urinary cfDNA. **a)** Large tumor with bladder infiltration showing strong molecular signals in urinary cfDNA. **b)** Intermediate-sized lesion without detectable molecular signals in plasma or urinary cfDNA. **c)** PI-RADS 3 lesion with a positive molecular signal in plasma cfDNA. **d)** Lesion with greater distance to the urethra demonstrating a molecular signal in plasma cfDNA, but not in urine. Yellow arrows highlight the index lesions. Green arrow depicts bladder infiltration (a).

### Multimodal correlation analyses revealed associations between radiological, serological, and cfDNA-based features in PCa patients

In order to investigate potential relationships between data modalities, correlation analyses were performed in all 33 PCa patients with complete datasets across radiomics, serolomics, and LBx features. Serological and radiological parameters included all features that had shown notable differences between PCa patients and cancer-free individuals in previous analyses, namely PSA, free PSA, free-to-total PSA ratio (free PSA %), PSA density, absolute monocyte count, androstenedione, and DHEA-S for serological markers, as well as ADC median, mean, minimum, 10^th^-percentile, ADC volume, and T2 volume for radiological features. LBx parameters involved all eight previously described plasma and urinary cfDNA features per patient. Additionally, for LBx markers assessed in both plasma and urine (TFx, CIA score, and methylation score), the maximum values across matched fluids were determined to capture the strongest signal per patient.

Among all PCa patients, several weak to moderate correlations emerged between radiological parameters and both LBx and serological features (**Figure 6**; *Supplementary Figure S6*), primarily involving the radiological features ADC minimum and ADC 10^th^-percentile, as well as ADC volume and T2 volume. Most correlations between radiomics and LBx features were inverse, in line with prior group-level findings where increasing tumor burden was associated with lower ADC values and elevated cfDNA-based tumor signals. Two exceptions were ADC or T2 volume and the plasma 10 bp-oscillation score. Both ADC and T2 volumes showed positive correlations with serological and LBx markers, while the plasma 10 bp-oscillation score, previously shown to decrease in PCa, demonstrated inverse associations with other tumor indicators. Due to strong internal correlations between ADC volume and T2 volume (ρ = 0.91; **Figure 6**; *Supplementary Figure S6*, *b*), results are reported representatively for ADC volume only. ADC volume correlated positively with plasma TFx (ρ = 0.38), the CIA score in plasma (ρ = 0.19), as well as the maximum values for CIA score (ρ = 0.23), TFx (ρ = 0.37) and methylation score (ρ = 0.34). Conversely, ADC minimum values showed negative correlations with multiple LBx metrics, including plasma TFx (ρ = –0.53), maximum TFx (ρ = –0.53), urinary CIA score (ρ = –0.26), maximum CIA score (ρ = –0.23), and urinary methylation score (ρ = –0.27), whereas a positive correlation was observed with the plasma 10 bp-oscillation score (ρ = 0.63). In line with its strong correlation with ADC minimum (ρ = 0.82) and the corresponding trends, ADC 10th-percentile additionally showed a negative association with the estimated TFx in urine (ρ = –0.27).

**Figure 6:**
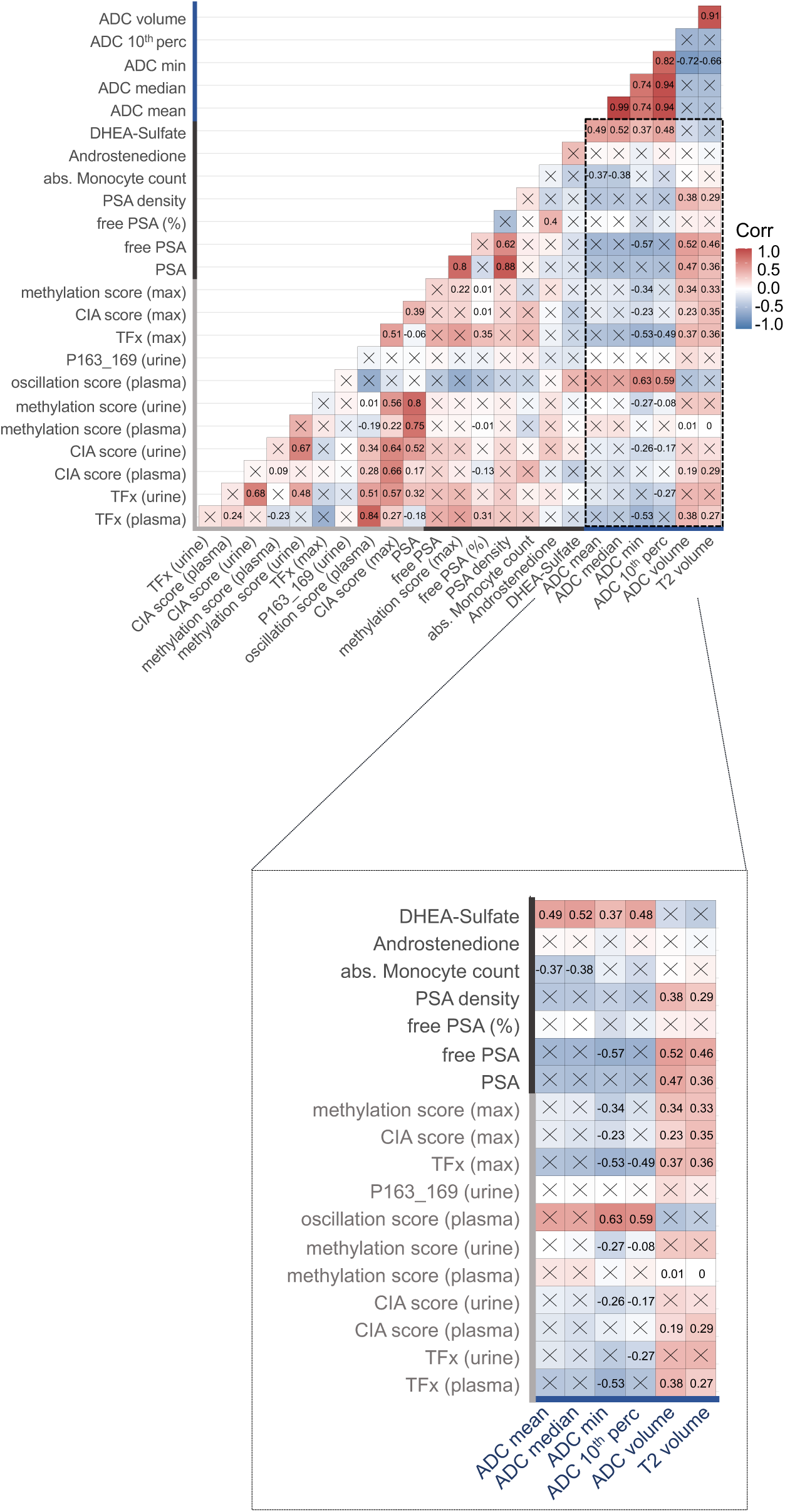
Correlation matrix of pairwise comparisons between relevant laboratory parameters, radiological features and liquid biopsy markers for all PCa patients. Positive correlations are shown in red and negative correlations in blue, with color intensity proportional to the correlation strength. Spearman correlation coefficients (Rho, ρ) are shown for significant correlations; statistically non-significant correlations are marked with an ‘x’.

With respect to serological parameters, ADC minimum also showed a negative correlation with free PSA levels (ρ = –0.57) and a positive correlation with DHEA-S (ρ = 0.39), while ADC volume correlated positively with PSA-related parameters, including free PSA (ρ = 0.52), total PSA (ρ = 0.47), and the PSA density (ρ = 0.38). Significant correlations with LBx features were exclusively observed for the free PSA%, particularly with the maximum TFx and plasma TFx (ρ = 0.35 and 0.31, respectively).

To summarize, multimodal correlation analysis revealed moderate associations between radiological, serological, and LBx parameters, with most relationships between radiomics and LBx, in line with prior observations linking increased tumor burden to reduced ADC values and elevated cfDNA signals.

## Discussion

This study investigated a comprehensive multimodal approach combining clinical parameters, serolomics, radiomics, and LBx markers for improved non-invasive detection and risk stratification of PCa. Several key findings emerged from the comprehensive assessment of all data layers. First, ctDNA was detectable in both plasma and urine samples from PCa patients in a real-world setting with detection rates notably improved through the integrative analysis of genomic and epigenomic features across both biofluids. This was particularly evident in cases with low PSA levels or localized disease, where conventional approaches show limited precision. Second, quantitative MRI-derived radiomic features and PSA-related serological parameters, alongside specific hormonal and immune cell features, revealed significant differences between PCa patients and cancer-free individuals, with many of these parameters also exhibiting gradual trends across disease stages. Many of the most informative radiological and serological parameters differentiated between lPCa and aPCa, aligning with prior evidence for their relevance in assessing both presence and aggressiveness of disease. Third, our study demonstrates that routine mpMRI, extended serological tests and complementary cfDNA analyses can be combined within a clinical screening workflow and reveal biologically meaningful associations. Radiomic features such as ADC minimum and 10^th^-percentile values correlated inversely with ctDNA levels captured by LBx metrics (TFx, CIA score, and methylation score), suggesting that reduced diffusion on MRI reflects increased tumor burden and greater genomic and epigenomic instability. Conversely, ADC and T2 lesion volumes correlated positively with PSA levels and LBx metrics, consistent with larger tumors releasing more ctDNA. These findings extend the concept of radiogenomics, linking imaging features to tissue-based genomic alterations, to circulating biomarkers and demonstrate that non-invasive imaging phenotypes carry systemic molecular information. The significant correlations of ADC-based radiomic features with cfDNA parameters and serological markers, respectively, underscore the potential of imaging to non-invasively capture tumor characteristics and to improve disease stratification.

These findings align with prior studies reporting significantly lower mean and minimum ADC values in malignant compared to benign lesions, and in lesions from high-risk-compared to low-risk PCa^34–36^. Both minimum and mean ADC values have been shown to assist in differentiating benign from malignant prostate lesions and in predicting clinically significant PCa^24,34^. The 10^th^-percentile ADC value was also correlated to the Gleason score, suggesting potential utility for PCa risk stratification^37^. In addition, significantly increased T2 and ADC volumes have been observed in PCa with extraprostatic extension^38^, consistent with PI-RADS criteria^30^. ADC volume correlates with Gleason score and histopathological tumor volume^39^, and post-prostatectomy analyses have further linked larger tumor volume with features of aggressiveness, systemic progression, and poorer outcomes^40,41^.

Our findings also align with literature that underscores the diagnostic relevance of PSA and its derivatives. Total PSA, PSA density, and the free-to-total PSA ratio all showed consistent associations with PCa status and stage, in line with clinical guidelines recommending their use for risk stratification^3,10^. Both PSA density and the free-to-total PSA ratio have been shown to improve PCa detection and risk assessment^42,43^, as well as to predict clinically significant PCa^44,45^.

Similarly, sex steroid hormones like DHEA-S and androstenedione demonstrated trends consistent with prior work from our group, involving a larger cohort of 578 men, where lower levels of androstenedione and DHEA-S, along with decreased free-to-total PSA ratio and elevated SHBG were significantly associated with aggressive PCa^19^. In this study, integrating DHEA-S, androstenedione, and SHBG with clinical parameters significantly improved the early prediction of aggressive PCa^19^. This multi-marker model outperformed PSA-based approaches and emphasized the utility of routine, cost-effective laboratory values for non-invasive risk stratification. These findings align with prior work showing that, although associations between blood levels of androgens, SHBG, or DHEA-S and PCa remain partly inconsistent, these markers have repeatedly been linked to PCa aggressiveness^46–49^.

A significant elevation in absolute monocyte counts was observed in our PCa cohort. This aligns with prior studies linking elevated monocyte levels to PCa risk and aggressiveness, and outlining the predictive value even in men with PSA < 10 ng/mL^50,51^. This supports the hypothesis that immune-related blood markers may complement hormonal and PSA-related parameters in PCa risk assessment.

With regard to the LBx analyses, our findings demonstrate that the complementary assessment of plasma and urinary cfDNA provides a more comprehensive tumor characterization than the analysis of either source alone. Integration of genomic and epigenomic analyses substantially improved ctDNA detectability compared to single-parameter analyses, yielding an overall detection rate of 50% in the entire PCa cohort. Consistent with previous reports, sensitivities were higher in advanced disease^52^, while detection in lPCa is known to be challenging^53,54^. Nonetheless, the complementary analysis achieved a ctDNA detection in 48% of localized cases, including patients with PSA levels < 10 ng/mL. The present findings are conceptually aligned with a limited number of investigations in PCa that have performed complementary plasma and urinary cfDNA assessments^55,56^, as well as with several studies in other malignancies demonstrating the potential of multimodal LBx approaches to enhance ctDNA detection^57–59^.

The comparative analysis of serolomics, radiomics, and LBx parameters revealed moderate correlations across modalities. The absence of strong correlations likely reflects the small sample size and predominance of lPCa cases, where tumor signals are typically lower. To our knowledge, this multiparametric framework—combining serological, radiological, and (epi)genomic liquid biopsy features—has not previously been reported for PCa detection and risk stratification, although the value of pairing individual modalities (e.g., radiogenomics) has been demonstrated in prior studies^26,60^. For example, Parry et al. correlated preoperative mpMRI data with molecular profiling of 43 benign and malign prostate tissue cores after prostatectomy, and additionally assessed cfDNA and genomic DNA from blood of five patients. They addressed the tumor heterogeneity within MRI-targeted lesions, but could not detect any alterations in cfDNA^61^. Morrison and colleagues combined radiological features from computed tomography imaging with genomic analyses of circulating tumor cells and plasma cfDNA in patients with metastatic castration-resistant PCa^62^. In PCa screening, the prospective, population-based Stockholm3-MRI trial demonstrated that combining a multivariable blood test (including genetic variants, protein biomarkers, and clinical risk factors) with MRI-targeted biopsy increased detection of clinically significant PCa and reduced overdetection of low-grade disease compared to traditional screening^63^.

From the clinical point of view, these multimodal strategies offer a promising framework for enhanced PCa diagnostics, enabling more accurate, minimally-invasive risk stratification and guiding decisions regarding prostate biopsy, or subsequent therapy management. The parameters assessed in this study are obtainable during standard diagnostic workflows, making their integration highly feasible within real-world clinical settings. From a practical perspective, LBx and laboratory assays are rapid, minimally invasive, and cost-effective, with cfDNA sequencing completed within 2–3 weeks and serological testing available within 1–2 days. Radiological features can be extracted from routine prostate MRI without additional patient burden or procedures needed. Together, these modalities can provide a comprehensive, real-time assessment of tumor heterogeneity in a clinically accessible manner. Beyond their individual utility, integration of these complementary modalities holds promise for synergistic enhancement of diagnostic accuracy and clinical utility.

Recent work has shown that the integration of multiple data modalities improves the prediction of outcomes compared with traditional models based on single modalities. For instance, in a large real-world dataset spanning several cancer types, random survival forests that incorporated all available variables (including clinical, genomic, pathology and natural language processing features) outperformed models based solely on tumor stage and prior progression^64^. Our study aligns with this concept by demonstrating that even within a relatively small cohort, combining radiological, serological and cfDNA markers revealed richer biological signals. This integration of multiple data modalities could provide a holistic view of tumor biology, enabling personalized risk assessment, as well as guiding biopsy and treatment decisions. By embedding multimodal data collection into routine clinical workflows, the ability to capture comprehensive, orthogonal data from every patient will be pivotal for developing predictive models that better reflect the diversity of patients seen in daily practice. Furthermore, these models could be embedded into digital twin frameworks, allowing continuous updates of individual patient risk profiles over time and supporting adaptive management strategies. By capturing radiological, serological and cfDNA features simultaneously, our dataset strongly contributes toward the development of patient-specific digital twins for PCa.

The present study relies on real world data (RWD) gathered from a prospectively enrolled screening cohort rather than a highly selected clinical trial population. RWD analyses are increasingly recognized for their ability to complement randomized controlled trials and to describe the reliability and transferability of complex procedures, such as molecular diagnostics and multimodal treatments, in routine clinical practice^65^. However, RWD studies often suffer from small sample sizes, making it challenging to extract robust biological insights. We addressed this limitation by generating multidimensional data for each participant. This strategy maximizes the information gained per patient and can mitigate the scarcity of real-world cohorts. Future research should focus on validating these findings in larger cohorts and in prospective clinical trials, to determine their impact on patient outcomes, cost-effectiveness, and integration into existing clinical workflows. With further validation, multimodal diagnostic models could not only enhance PCa management but also be adapted to other malignancies, contributing to the broader vision of precision oncology.

A major limitation of this study was the small sample size, particularly for the control group and aPCa subgroup, which limited statistical power and generalizability. Moreover, the integration of different data modalities required complete datasets, further reducing the analyzable cohort. The small sample size did not allow for the intended development of a robust machine learning–based, multiparametric prediction model for PCa risk stratification. While radiological feature extraction holds considerable promise, the interpretation of ADC values can be influenced by technical factors such as field strength and b-values, highlighting the need for standardization and harmonization across imaging protocols^66,67^. Similarly, radiological analyses were limited to first-order features without standardization of signal intensities in T2-weighted imaging, which may have introduced variability^68,69^. In addition, the absence of an independent validation cohort restricts the generalizability of the findings and precludes assessment of their robustness in external populations.

In conclusion, we demonstrate in this proof-of-principle study that the integration of radiomics, serolomics, and LBx analyses captures complementary tumor-related information and improves the potential for non-invasive detection and risk stratification in a real-world PCa screening cohort. Multimodal assessment can complement established diagnostic workflows, particularly in cases where single-modality results are inconclusive. The use of routinely obtainable laboratory parameters, standard imaging protocols, and increasingly accessible cfDNA assays supports the feasibility of this holistic non-invasive approach for future clinical implementation. With further validation in larger, prospective cohorts, integrated multiparametric models could enhance precision oncology in PCa and other tumor entities.

## Methods

### Study patients

Forty-nine men who underwent PCa screening were recruited at Heidelberg University Hospital (June 2021 – November 2022). All individuals provided informed consent and the study was approved by the ethic committee of the Medical Faculty of Heidelberg University (S-130/2021). The diagnostic procedure involved routine blood collection and laboratory assessment, radiological assessment of the prostate with mpMRI, and imaging-guided prostate tissue biopsy with subsequent histopathological assessment. Comprehensive clinical data was prospectively collected for all men. Further information regarding family history (first-degree relatives with PCa), lifestyle factors (diet, alcohol consumption, smoking and physical examination), and other potential risk factors for PCa (infertility, sterilization, occurrence of urinary tract infection or urinary retention within the last 6 months) were assessed via questionnaire.

This cohort represents a subset of a larger prospective dataset comprising 578 men who underwent diagnostic evaluation for suspected PCa^19^. In addition, LBx data were derived from a previously published study involving a larger cohort of PCa patients and cancer-free individuals^31^. The present analysis was restricted to participants with available LBx, serological, radiological, and survey data, allowing for integrative correlation analyses across all data layers.

MpMRI of the prostate was performed at the Radiological department of the German Cancer Research Center (DKFZ) Heidelberg, and was assessed following the PI-RADS recommendations^30^. Prostate tissue biopsies at Urology Clinic Heidelberg were performed as transperineal fusion-targeted biopsy of PI-RADS 3–5 lesions identified in the prostate MRI, with elastic software registration using a UroNav system (Philips Invivo, Gainsville, FL, USA), in addition to systematic, saturation biopsy adjusted to prostate volume, as previously described^70,71^. Histopathological assessment of the prostate tissue biopsies was performed at the Institute of Pathology of Heidelberg University Hospital, according to International Society of Urological Pathology standards^72^.

### Assessment of routine laboratory parameters

Routine laboratory assessments were conducted for all men, including small blood count, coagulation parameters, as well as renal and liver function parameters, amongst others. PCa screening also included the assessment of total PSA levels. The routine laboratory assessments were complemented by additional 31 parameters, involving inflammation, metabolism, hormones, vitamins and prostate-related markers (*Supplementary Table S4*). The internal laboratory blood assessments were performed by the central laboratory of Heidelberg University Hospital, which has been accredited according to DIN EN ISO 15189 since 2005. The extended serological parameters were previously investigated in a larger prospective cohort of men undergoing diagnostic evaluation for suspected PCa, with systematic assessment of their associations to clinical and pathological features, including disease aggressiveness^19^.

### Assessment of radiological features derived from quantitative analysis of mpMRI

MpMRI was conducted at the Radiological department of the DKFZ Heidelberg, as previously reported^73^. The mpMRI was performed using the 3.0 Tesla scanner (Magnetom Prisma, Biograph mMR; Siemens Healthineers) for majority of the patients, and in few cases the 1.5 Tesla MRI scanner (Magnetom Aera, Siemens Healthineers). The radiological assessment was based on PI-RADS recommendations^30,74^ and guidelines of the European Society of Urogenital Radiology^75,76^. It was performed during clinical routine by experienced radiologists with access to clinical data, and confirmed by board-certified radiologists. Suspicious prostate lesions were reported, according to PI-RADS v2.1 guidelines^74^, and were subsequently subjected to transperineal fusion-targeted biopsy. First-order parameters (minimum, maximum, median, mean, 10^th^-percentile, 90^th^-percentile, standard deviation, variance, volume) from axial T2-weighted images (T2w) and apparent diffusion coefficient (ADC) maps, respectively, were extracted for the manually segmented, suspicious lesions. For retrospective comparison of the radiological parameters with LBx and serolomics data, several targeted lesions were reduced to one representative radiological index lesion for each man. These lesions were defined by a radiologist experienced in the assessment of prostate MRIs who was blinded for the histopathological result.

### Genomic and epigenomic LBx analyses

Additional blood (2 x 9 ml) and urine (30-50ml) for LBx analyses were collected prior to prostate biopsy for the majority of men. In case of nine men, samples were collected at the follow-up visit after the tissue biopsy, but with sufficient time interval to avoid post-procedural effects on the LBx analyses. CfDNA was isolated from plasma and urine, and genome-wide methylation profiling with cell-free methylated DNA immunoprecipitation sequencing and low-coverage whole genome sequencing were performed following established protocols^31^. Genome-wide methylation analysis, CNV profiling, chromosomal instability assessment, and cfDNA fragmentation analysis were conducted, as described previously^31^.

For each participant, a total of eight genomic and epigenomic cfDNA features were assessed for ctDNA detection in plasma and urine: the estimated TFx from the CNV analysis, the CIA score, the methylation score, and two cfDNA fragmentation features for plasma and urine (10 bp-oscillation score in plasma; proportion of cfDNA fragments with 163–169 bp lengths (P163–169 bp) in urine). TFx, CIA score, and methylation score were evaluated in both plasma and urinary cfDNA. Previously established detection thresholds based on the 95^th^-percentile values (TFx, CIA score, methylation score, P163–169 bp in urine) or the 5^th^-percentile values (10bp-oscillation in plasma) of the cancer-free control cohort for these features were applied to classify samples as ctDNA-positive^31^.

### Statistical analyses and data visualization

All statistical analyses were performed in R v4.0.0^77^. Group comparisons were conducted using the Kruskal-Wallis test with Dunn’s post hoc test, Wilcoxon rank-sum test, or Fisher’s exact test, as appropriate. P values were adjusted for multiple testing with Benjamini-Hochberg‘s method, with significance set at adjusted p < 0.05. Visualizations were generated using R package ggplot2^78^.

Pairwise correlations between features were assessed using Spearman’s rank correlation. Prior to analysis, variables were standardized using z-score transformation. The cor.test() function from R package stats v.4.0.0^77^ was used to compute Spearman’s correlation coefficients (ρ) and corresponding p values. Correlation matrices were visualized using the R package ggcorrplot (v0.1.4.1) ^79^, where positive correlations were shown in red, negative in blue, and non-significant correlations were either blank or marked with a cross. Correlation strength was interpreted as follows: 0.00–0.19 (very weak), 0.20–0.39 (weak), 0.40–0.59 (moderate), 0.60–0.79 (strong), and 0.80–1.00 (very strong).

Correlation analyses included LBx-derived features, serological markers, and radiological parameters. For the LBx data, all plasma and urinary cfDNA features described above were included. Additionally, composite scores were generated by selecting the maximum value between matched plasma and urine for TFx, CIA score, and methylation score, resulting in a total of eleven LBx-derived features per patient for correlation analyses. For laboratory and imaging features, only those previously identified as significantly different between PCa patients and cancer-free individuals were included.

### Usage of generative Artificial Intelligence (AI) and AI-assisted technologies

During the preparation of this work, the authors used ChatGPT-4o in order to improve language and readability. After using this tool, the authors reviewed and edited the content as needed, and take full responsibility for the content of the publication.

## Data availability

The data sets generated and analyzed during the current study are available from the corresponding author on reasonable request.

## Code availability

All R packages and bioinformatic tools used in this study are publicly available, as described in the Methods section. Any additional information, including custom code for data processing and visualization, required to reanalyze the data reported in this work, is available from the corresponding author upon reasonable request.

## Acknowledgements

The authors thank all members of the Cancer Genome Research Group at German Cancer Research Center (DKFZ) for their support and constructive discussions. We also acknowledge the Division of Radiology at the DKFZ for their excellent collaboration, which made this interdisciplinary work possible. In addition, we are grateful to the DKFZ core facilities for their assistance, in particular the NGS Core Facility for sequencing analyses and the Omics IT and Data Management Core Facility for data management and processing. We sincerely acknowledge our patients and their caregivers, as well as the help and support of the (medical) staff at the Urology Clinic of Heidelberg University Hospital.

This work was realized through support by the German Federal Ministry for Economic Affairs and Climate Action (funding # 01MT21004A) and the Dieter Morszeck Foundation. The funders had no role in the design of the study; in the collection, analyses, or interpretation of data; in the writing of the manuscript, or in the decision to publish the results.

## Contributorship

Conceptualization: A.L.R., M.G., C.M., D.B.; Data curation: A.L.R., S.E., D.J., O.L., C.M.; Formal analysis: A.L.R., C.M., O.L.; Funding acquisition: M.G.; Investigation: M.G., D.B., A.L.R., C.M.; Methodology: H.S., M.G., A.L.R., C.M., F.J.; Project Administration: M.G.; Resources: M.G., A.L.R.; Software: A.L.R., O.L., J.F.; Statistical analysis: A.L.R., C.M., O.L.; Supervision: M.G., H.S., D.H., O.S., S.F., D.B., H.P.S.; Visualization: A.L.R., C.M.; Writing - original draft: A.L.R., M.G.; Writing - Review & Editing: C.M., D.J., S.E., F.J., O.L., D.H., O.S., S.F., D.B., H.P.S., H.S., M.G. All authors critically reviewed and approved the final version of the manuscript.

## Ethical approval

The study was approved by the ethical committee of the University of Heidelberg (Approval No. S-130/2021) and was performed in accordance with the Declaration of Helsinki. Written informed consent was obtained from all participants prior to study inclusion.

