## Supplementary Material for "Combined radiomics and liquid biopsy reflect tumor biology towards multimodal non-invasive prostate cancer risk stratification"

### Supplementary Tables

Table S1: Overview of results from clinical and survey data, assessing potential PCa risk and lifestyle factors

Table S2: Assessment of first-order radiological features from prostate MRI based on T2-weighted imaging or ADC mapping for 42 index lesions of 23 PCa patients and 9 cancer-free individuals

Table S3: Statistical comparison of first-order radiological features from prostate MRI between cancer-free individuals, localized and advanced PCa patients

Table S4: Assessment of 31 laboratory parameters in the blood from 36 PCa patients and 13 cancer-free individuals

Table S5: Statistical comparison of 31 laboratory parameters between cancer-free individuals, localized and advanced PCa patients

### Supplementary Figures

Figure S1: Assessment of first-order radiological features from prostate MRI based on T2-weighted imaging or ADC mapping for 42 index lesions of 33 PCa patients and 9 cancer-free individuals

Figure S2: Assessment of prostate-associated parameters measured in the blood from PCa patients and cancer-free individuals

Figure S3: Assessment of sex and adrenal hormones measured in the blood from PCa patients and cancer-free individuals

Figure S4: Assessment of differential blood count and inflammation parameters measured in the blood from PCa patients and cancer-free individuals

Figure S5: Assessment of metabolic parameters and vitamins measured in the blood from PCa patients and cancer-free individuals

Figure S6: Spearman correlation matrices of radiological features, serological parameters, and liquid biopsy markers in all PCa patients

### Supplementary Tables

**Supplementary Table S1:** Overview of results from clinical and survey data, assessing potential PCa risk and lifestyle factors

| **clinical parameter** | **control** median, range | **tumor** median, range | **tumor vs. control** p-value [adjusted p-value] |
|---|---|---|---|
| **age** | 60 [57 ; 74] | 66 [49 ; 79] | 0.061 [0.265] |
| **meat consumption** (days/week) | 3 [1 ; 7] NA: 0 | 3 [0 ; 7] NA: 3 | 0.627 [1.0] |
| **alcohol consumption** (days/week) | 2 [0 ; 6] NA: 0 | 2 [0 ; 7] NA: 5 | 0.989 [1.0] |
| **physical examination** (days/week) | 4 [0 ; 10] NA: 0 | 3 [0 ; 18] NA: 5 | 0.459 [0.862] |
| **smoking** (packyears) | 0 [0 ; 24] NA: 0 | 0 [0 ; 0] NA: 5 | 0.030 [0.265] |
| **body mass index** (bmi; kg/m$^2$) | 26.7 [18.7 ; 35.5] NA: 0 | 26.2 [21.0 ; 42.1] NA: 2 | 1.0 [1.0] |
| **PCa family history** count (yes/no) | 2 ; 11 NA: 0 | 7 ; 27 NA: 2 | 1.0 [1.0] |
| **current smoker** count (yes/no) | 2 ; 11 NA: 0 | 1 ; 35 NA: 0 | 0.168 [0.546] |
| **infertility** count (yes/no) | 1 ; 12 NA: 0 | 1 ; 35 NA: 0 | 0.464 [0.862] |
| **sterilization** count (yes/no) | 1 ; 12 NA: 0 | 2 ; 34 NA: 0 | 1.0 [1.0] |
| **urinary tract infection (last 6 months)** count (yes/no) | 1 ; 12 NA: 0 | 4 ; 32 NA: 0 | 1.0 [1.0] |
| **urinary retention (last 6 months)** count (yes/no) | 0 ; 13 NA: 0 | 5 ; 31 NA: 0 | 0.306 [0.797] |
| **digital rectal examination, suspicious finding** count (yes/no) | 0 ; 13 NA: 0 | 10 ; 26 NA: 0 | 0.045 [0.265] |

Median with range for discrete variables (top) and number of men with positive/negative answer for categorical variables (bottom) for cancer-free individuals and PCa patients. Last column: Statistical comparison between results from PCa patients and cancer-free individuals. Significance testing for the comparison of discrete variables with Wilcoxon rank test, and for categorical variables with fisher-test. Adjustment for multiple testing with Benjamini-Hochberg, adjusted p value $< 0.05$. Adjusted $p < 0.05$ was defined as statistically significant results. PCa patients, n = 36; cancer-free individuals, n = 13. Complete data sets for 40 men (PCa patients, n = 27; cancer-free individuals, n = 13). Abbreviations: NA = data not available, PCa = prostate cancer

**Supplementary Table S2:** Assessment of first-order radiological features from prostate MRI based on T2-weighted imaging or ADC mapping for 42 index lesions of 33 PCa patients and 9 cancer-free individuals

| **radiological feature** | **control** median, range | **tumor** median, range | **localized PCa** median, range | **advanced PCa** median, range | **tumor vs. control** p value [adjusted p value] |
|---|---|---|---|---|---|
| **ADC 10th-percentile** | 809 [565 ; 1246] | 582 [232 ; 936] | 593 [232 ; 936] | 518 [257 ; 657] | **0.01 [0.036]** |
| **ADC 90th-percentile** | 1084 [891 ; 1546] | 930 [682 ; 1455] | 930 [682 ; 1455] | 933 [730 ; 1058] | 0.023 [0.063] |
| **ADC maximum** | 1234 [935 ; 1684] | 1176 [823 ; 1973] | 1142 [823 ; 1629] | 1287 [828 ; 1973] | 0.366 [0.434] |
| **ADC mean** | 924 [746; 1394] | 737 [486 ; 1160] | 755 [486 ; 1160] | 654 [563 ; 852] | **0.01 [0.036]** |
| **ADC median** | 914 [732 ; 1427] | 740 [461 ; 1160] | 740 [461 ; 1160] | 643 [559 ; 841] | **0.011 [0.036]** |
| **ADC minimum** | 693 [362 ; 991] | 426 [14 ; 882] | 460 [22 ; 882] | 74 [14 ; 532] | **0.005 [0.034]** |
| **ADC SD** | 141 [92 ; 217] | 141 [46 ; 281] | 132 [46 ; 233] | 157 [77 ; 281] | 0.786 [0.786] |
| **ADC variance** | 20010 [8401 ; 47218] | 19940 [2116 ; 79130] | 17433 [2116 ; 5406] | 24785 [5927 ; 79130] | 0.786 [0.786] |
| **ADC volume** | 0.24 [0.13 ; 1.13] | 1.24 [0.02 ; 66.3] | 1.21 [0.02 ; 7.19] | 8.43 [0.68 ; 66.38] | **0.002 [0.023]** |
| **T2 10th-percentile** | 156 [48 ; 264] | 128 [65 ; 224] | 130 [65 ; 224] | 92.5 [78 ; 147] | 0.249 [0.348] |
| **T2 90th-percentile** | 326 [179 ; 725] | 245 [161 ; 414] | 264 [161 ; 414] | 216 [179 ; 255] | 0.078 [0.135] |
| **T2 maximum** | 488 [338 ; 969] | 471 [251 ; 869] | 474 [251 ; 839] | 378 [322 ; 869] | 0.635 [0.709] |
| **T2 mean** | 255 [126 ; 355] | 184 [111 ; 317] | 197 [111 ; 317] | 153 [130 ; 197] | 0.049 [0.105] |
| **T2 median** | 253 [123 ; 343] | 184 [108 ; 314] | 192 [108 ; 314] | 151 [127 ; 199] | 0.05 [0.105] |
| **T2 minimum** | 72 [4 ; 149] | 18 [1 ; 165] | 25 [1 ; 165] | 4 [1 ; 62] | 0.101 [0.160] |
| **T2 SD** | 55 [40 ; 267] | 52 [28 ; 91] | 53 [28 ; 91] | 46 [38 ; 57] | 0.275 [0.348] |
| **T2 variance** | 3042 [1570 ; 71069] | 2659 [810 ; 8277] | 2842 [810 ; 8277] | 2130 [1424 ; 3210] | 0.275 [0.348] |
| **T2 volume** | 0.27 [0.15 ; 0.93] | 1.61 [0.07 ; 57.16] | 1.59 [0.07 ; 7.65] | 7.77 [0.43 ; 57.16] | **0.0004 [0.009]** |

Units: ADC values: $mm^2/s$; T2 values: ms; ADC/T2 volume: ml; Median with range of each radiological feature for all PCa patients, cancer-free individuals, as well as tumor patients stratified for localized and advanced PCa. Last column: statistical comparison between all PCa patients and cancer-free individuals. Significance testing with Wilcoxon rank sum test; multiple testing with Benjamini Hochberg, significant values with adjusted p value < 0.05; significant results highlighted in bold letters; PCa patients, n = 33; cancer-free individuals, n = 9. Abbreviations: ADC = apparent diffusion coefficient, MRI = magnetic resonance imaging, SD = standard deviation

**Supplementary Table S3:** Statistical comparison of first-order radiological features from prostate MRI between cancer-free individuals, localized and advanced PCa patients

| radiological feature | kruskal wallis test<br>control vs. localized PCa vs. advanced PCa p value | Dunn´s Post Hoc Test<br>control vs. localized PCa p value [adjusted p value] | control vs. advanced PCa p value [adjusted p value] | localized PCa vs. advanced PCa p value [adjusted p value] |
|---|---|---|---|---|
| **ADC 10$^{th}$-percentile** | **0.008** | 0.038 [0.056] | **0.002 [0.006]** | 0.066 [0.066] |
| **ADC 90$^{th}$-percentile** | 0.066 | 0.038 [0.063] | 0.042 [0.063] | 0.541 [0.541] |
| **ADC maximum** | 0.364 | 0.259 [0.418] | 0.918 [0.918] | 0.278 [0.418] |
| **ADC mean** | **0.01** | 0.034 [0.051] | **0.003 [0.009]** | 0.098 [0.098] |
| **ADC median** | **0.009** | 0.034 [0.052] | **0.002 [0.007]** | 0.081 [0.081] |
| **ADC minimum** | **0.003** | **0.026 [0.04]** | **0.001 [0.002]** | **0.043 [0.043]** |
| **ADC SD** | 0.32 | 0.975 [0.975] | 0.213 [0.319] | 0.138 [0.319] |
| **ADC variance** | 0.32 | 0.975 [0.975] | 0.213 [0.319] | 0.138 [0.319] |
| **ADC volume** | **0.002** | **0.014 [0.021]** | **0.001 [0.002]** | 0.057 [0.057] |
| **T2 10$^{th}$-percentile** | 0.143 | 0.415 [0.415] | 0.051 [0.153] | 0.114 [0.17] |
| **T2 90$^{th}$-percentile** | **0.049** | 0.169 [0.169] | **0.014 [0.042]** | 0.091 [0.136] |
| **T2 maximum** | 0.833 | 0.686 [0.724] | 0.55 [0.724] | 0.724 [0.724] |
| **T2 mean** | **0.04** | 0.111 [0.111] | **0.012 [0.035]** | 0.111 [0.111] |
| **T2 median** | **0.04** | 0.111 [0.111] | **0.012 [0.035]** | 0.111 [0.111] |
| **T2 minimum** | **0.021** | 0.254 [0.254] | **0.006 [0.018]** | **0.026 [0.038]** |
| **T2 SD** | 0.198 | 0.428 [0.428] | 0.074 [0.222] | 0.158 [0.237] |
| **T2 variance** | 0.198 | 0.428 [0.428] | 0.074 [0.222] | 0.158 [0.237] |
| **T2 volume** | **0.001** | **0.004 [0.006]** | **0.0004 [0.001]** | 0.087 [0.087] |

Significance testing for multiple comparisons with Kruskal wallis Test, $p < 0.05$ for significant results; Post Hoc Test with Dunn´s Test, multiple testing with Benjamini-Hochberg, significant values with adjusted $p < 0.05$; significant results highlighted in bold letters. Localized PCa patients, n = 27; advanced PCa patients, n = 6; cancer-free individuals, n = 9.

**Supplementary Table S4:** Assessment of 31 laboratory parameters in the blood from 36 PCa patients and 13 cancer-free individuals

| **laboratory parameter** [unit, reference] | **control** median, range | **tumor** median, range | **localized PCa** median, range | **advanced PCa** median, range | **tumor vs. control** p value [adjusted p value] |
|---|---|---|---|---|---|
| **Leukocytes** 4-10 /nL | 5.9 [4.4 ; 9.7] | 6.7 [3.5 ; 13.3] | 6.7 [3.5 ; 11.9] | 7.4 [4.7 ; 13.3] | 0.063 [0.252] |
| **Hemoglobin** 13-17 g/dL | 15.2 [13.3 ; 17.2] | 15.5 [7.2 ; 18.4] | 15.56 [12.3 ; 18.4] | 14.8 [7.2 ; 16.2] | 0.437 [0.669] |
| **Platelets** 150-440 /nL | 244 [151 ; 402] | 249 [149 ; 344] | 249 [149 ; 344] | 257 [215 ; 315] | 0.668 [0.841] |
| **Neutrophils (%)** 50-80 % | 65.1 [49.4 ; 83.6] | 65.3 [49.9 ; 78.2] | 65.1 [49.9 ; 78.2] | 65.7 [56.2 ; 77.1] | 0.981 [0.981] |
| **Lymphocytes (%)** 25-40 % | 24.4 [10.5 ; 40.1] | 23.7 [12.4 ; 33.7] | 23.7 [13.5 ; 33.7] | 22.7 [12.4 ; 29.4] | 0.981 [0.981] |
| **Monocytes (%)** 2-9% | 5.3 [4 ; 7.1] | 6.3 [3.9 ; 8.8] | 6.2 [3.9 ; 8.8] | 6.7 [4.2 ; 8.4] | **0.033** [0.188] |
| **Absolute Neutrophil Count** 1.8-7.7 /nL | 3.5 [2.5 ; 8.1] | 4.3 [2.0 ; 9.8] | 4.3 [2 ; 9.0] | 4.9 [2.7 ; 9.8] | 0.122 [0.297] |
| **Absolute Lymphocyte Count** 1.0-4.8 /nL | 1.4 [0.95 ; 2.4] | 1.6 [1.2 ; 3.4] | 1.6 [1.2 ; 3.4] | 1.4 [1.2 ; 2.2] | 0.085 [0.262] |
| **Absolute Monocyte Count** 0.2-0.8 /nL | 0.3 [0.2 ; 0.4] | 0.4 [0.2 ; 0.9] | 0.4 [0.2 ; 0.9] | 0.4 [0.3 ; 0.9] | **0.006** [0.064] |
| **C-Reactive Protein** <5 mg/L | 2 [2 ; 48.2] | 2 [2 ; 26.3] | 2 [2 ; 24] | 2 [2 ; 26.3] | 0.917 [0.975] |
| **Progesterone** <0.16-0.48 nmol/L | 0.4 [0.1 ; 0.7] | 0.3 [0.1 ; 0.7] | 0.4 [0.1 ; 0.7] | 0.3 [0.2 ; 0.5] | 0.071 [0.252] |
| **Estradiol/E2** 10-45 pg/mL | 27 [17 ; 40] | 30 [14 ; 48] | 30 [14 ; 42] | 30 [24 ; 48] | 0.074 [0.252] |
| **Testosterone** 2-7 ng/mL | 5 [2 ; 9] | 4 [2 ; 7] | 4 [2 ; 7] | 4 [2 ; 7] | 0.372 [0.602] |
| **Androstenedione** 75-275 ng/dL | 94 [48 ; 151] | 66.5 [18 ; 120] | 66 [18 ; 120] | 68 [54 ; 117] | **0.008** [0.064] |
| **Dihydrotestosterone** 25-115 ng/dL | 40 [13 ; 86] | 31 [2 ; 82] | 31 [14 ; 82] | 26 [2 ; 81] | 0.319 [0.542] |
| **DHEA-Sulfate** xx µg/mL (age-related differences) | 1.4 [0.7 ; 3.2] | 0.8 [0.2 ; 3.2] | 0.9 [0.2 ; 3.2] | 0.6 [0.5 ; 1.5] | **0.006** [0.064] |
| **Sex Hormone Binding Globulin** 10-72 nmol/L | 42 [22 ; 67] | 44 [19 ; 91] | 43 [20 ; 87] | 58 [19 ; 91] | 0.546 [0.714] |
| **Cortisol** 56-200 ng/mL | 165 [107 ; 271] | 158 [70 ; 273] | 156 [70 ; 273] | 194 [98 ; 242] | 0.472 [0.669] |
| **PSA** <4 ng/mL | 5.6 [4.2 ; 14.4] | 8.3 [1.8 ; 391.3] | 7.9 [1.8 ; 28.7] | 29.7 [6.3 ; 391.3] | 0.116 [0.297] |
| **free PSA** xx ng/mL | 1.3 [0.5 ; 3.6] | 1.5 [0.3 ; 37.0] | 1.2 [0.3 ; 2.7] | 2.5 [1.5 ; 37.0] | 0.883 [0.975] |
| **free PSA (%)** >15 % | 24 [6 ; 50] | 15.6 [7 ; 43] | 15 [7 ; 39] | 22 [8 ; 43] | **0.009** [0.064] |

| | | | | | |
|---|---|---|---|---|---|
| **PSA density**<br>xx ng/mL$^2$ | 0.09<br>[0.03 ; 0.22] | 0.19<br>[0.06 ; 4.01] | 0.18<br>[0.06 ; 0.95] | 0.36<br>[0.07 ; 4.01] | **0.001** [**0.027**] |
| **Vitamin A**<br>1.05-2.8 µmol/L | 2.4<br>[1.5 ; 3.3] | 2.2<br>[1.2 ; 4.3] | 2.25<br>[1.5 ; 4.3] | 2.1<br>[1.2 ; 2.7] | 0.545 [0.714] |
| **Vitamin E**<br>12-42 µmol/L | 45<br>[34 ; 61] | 42<br>[21 ; 66] | 42<br>[21 ; 66] | 40<br>[28 ; 58] | 0.093 [0.262] |
| **Zinc**<br>70-150 µg/dL | 12<br>[9 ; 14] | 12<br>[9 ; 15] | 12<br>[9 ; 15] | 12<br>[9 ; 13] | 0.816 [0.975] |
| **25-Hydroxy Vitamin D**<br>6.3-46.4 ng/mL | 19.0<br>[8.1 ; 49.5] | 21.9<br>[10.1 ; 102.9] | 20.9<br>[10.1 ; 80.3] | 24.3<br>[10.4 ; 102.9] | 0.862 [0.975] |
| **Glucose**<br>65- 110 mg/dL | 102<br>[76 ; 141] | 102<br>[59 ; 273] | 102<br>[59 ; 273] | 92<br>[83 ; 129] | 0.898 [0.975] |
| **HbA1c (IFCC)**<br>20-42 mmol/mol | 36<br>[31 ; 43] | 38<br>[30 ; 66] | 38<br>[30 ; 66] | 36<br>[30 ; 48] | 0.170 [0.339] |
| **Alkaline Phosphatase**<br>40-130 U/L | 83<br>[59 ; 132] | 71<br>[49 ; 229] | 71<br>[49 ; 103] | 71<br>[51 ; 229] | 0.210 [0.397] |
| **Triglycerides**<br><150 mg/dL | 152<br>[64 ; 240] | 103<br>[33 ; 297] | 115<br>[33 ; 297] | 75<br>[59 ; 173] | 0.234 [0.419] |
| **HDL Cholesterol**<br>>40 mg/dL | 45<br>[32 ; 70] | 45<br>[31 ; 85] | 47<br>[31 ; 85] | 43<br>[34 ; 59] | 0.465 [0.669] |

Median with range of each laboratory parameter for all PCa patients, cancer-free individuals, as well as tumor patients stratified for localized and advanced PCa. Last column: statistical comparison between all PCa patients and cancer-free individuals. Significance testing with Wilcoxon rank sum test; multiple testing with Benjamini Hochberg, significant values with adjusted p value < 0.05; significant results highlighted in bold letters; PCa patients, n = 36; cancer-free individuals, n = 13. Abbreviations: DHEA = Dehydroepiandrosterone, HbA1c = Hemoglobin A1c, HDL = high-density lipoprotein, IFCC = International Federation of Clinical Chemistry and Laboratory Medicine, PSA = prostate-specific antigen

**Supplementary Table S5:** Statistical comparison of 31 laboratory parameters between cancer-free individuals, localized and advanced PCa patients

| laboratory parameter | kruskal wallis test | Dunn´s Post Hoc Test | | |
|---|---|---|---|---|
| | control vs. localized PCa vs. advanced PCa p value | control vs. localized PCa p value [adjusted p value] | control vs. advanced PCa p value [adjusted p value] | localized PCa vs. advanced PCa p value [adjusted p value] |
| **Leukocytes** | 0.175 | 0.072 [0.217] | 0.185 [0.278] | 0.966 [0.966] |
| **Hemoglobin** | 0.210 | 0.245 [0.368] | 0.554 [0.554] | 0.114 [0.343] |
| **Platelets** | 0.793 | 0.767 [0.767] | 0.497 [0.767] | 0.604 [0.767] |
| **Neutrophils (%)** | 0.523 | 0.742 [0.742] | 0.428 [0.641] | 0.255 [0.641] |
| **Lymphocytes (%)** | 0.515 | 0.792 [0.792] | 0.394 [0.592] | 0.250 [0.592] |
| **Monocytes (%)** | 0.086 | 0.055 [0.086] | 0.057 [0.086] | 0.567 [0.567] |
| **Abs. Neutrophil Count** | 0.279 | 0.158 [0.271] | 0.180 [0.271] | 0.722 [0.722] |
| **Abs. Lymphocyte Count** | 0.160 | 0.059 [0.178] | 0.529 [0.529] | 0.421 [0.529] |
| **Abs. Monocyte Count** | **0.022** | **0.009** [**0.026**] | **0.043** [0.065] | 0.886 [0.886] |
| **C-Reactive Protein** | 0.802 | 0.781 [0.781] | 0.696 [0.781] | 0.513 [0.781] |
| **Progesterone** | 0.175 | 0.099 [0.175] | 0.117 [0.175] | 0.668 [0.668] |
| **Estradiol/E2** | 0.117 | 0.138 [0.208] | 0.047 [0.141] | 0.305 [0.305] |
| **Testosterone** | 0.660 | 0.397 [0.719] | 0.479 [0.719] | 0.911 [0.911] |
| **Androstenedione** | **0.026** | **0.007** [**0.022**] | 0.116 [0.175] | 0.703 [0.703] |
| **Dihydrotestosterone** | 0.470 | 0.421 [0.482] | 0.228 [0.482] | 0.482 [0.482] |
| **DHEA-Sulfate** | **0.018** | **0.010** [**0.031**] | **0.021** [**0.031**] | 0.597 [0.597] |
| **Sex Hormone Binding Globulin** | 0.455 | 0.749 [0.749] | 0.225 [0.411] | 0.274 [0.411] |
| **Cortisol** | 0.420 | 0.326 [0.488] | 0.777 [0.777] | 0.273 [0.488] |
| **PSA** | **0.008** | 0.377 [0.377] | **0.003** [**0.008**] | **0.008** [**0.012**] |
| **free PSA** | **0.002** | 0.316 [0.316] | **0.017** [**0.025**] | **0.001** [**0.002**] |
| **free PSA (%)** | **0.012** | **0.004** [**0.012**] | 0.443 [0.443] | 0.154 [0.230] |
| **PSA density** | **0.002** | **0.005** [**0.008**] | **0.001** [**0.004**] | 0.164 [0.164] |
| **Vitamin A** | 0.344 | 0.792 [0.792] | 0.167 [0.278] | 0.186 [0.278] |
| **Vitamin E** | 0.228 | 0.087 [0.261] | 0.335 [0.502] | 0.772 [0.772] |
| **Zinc** | 0.715 | 0.969 [0.969] | 0.464 [0.695] | 0.434 [0.695] |
| **25-Hydroxy Vitamin D** | 0.983 | 0.857 [0.998] | 0.899 [0.998] | 0.998 [0.998] |
| **Glucose** | 0.687 | 0.936 [0.936] | 0.476 [0.714] | 0.393 [0.714] |
| **HbA1c (IFCC)** | 0.249 | 0.114 [0.343] | 0.768 [0.768] | 0.352 [0.528] |
| **Alkaline Phosphatase** | 0.442 | 0.239 [0.469] | 0.313 [0.469] | 0.854 [0.854] |
| **Triglycerides** | 0.221 | 0.403 [0.403] | 0.082 [0.247] | 0.209 [0.314] |
| **HDL Cholesterol** | 0.585 | 0.368 [0.706] | 0.995 [0.995] | 0.470 [0.706] |

Significance testing for multiple comparisons with Kruskal wallis Test, $p < 0.05$ for significant results; Post Hoc Test with Dunn´s Test, multiple testing with Benjamini-Hochberg, significant values with adjusted $p < 0.05$; significant results highlighted in bold letters. Localized PCa patients, n = 29; advanced PCa patients, n = 7; cancer-free individuals, n = 13.

### Supplementary Figures

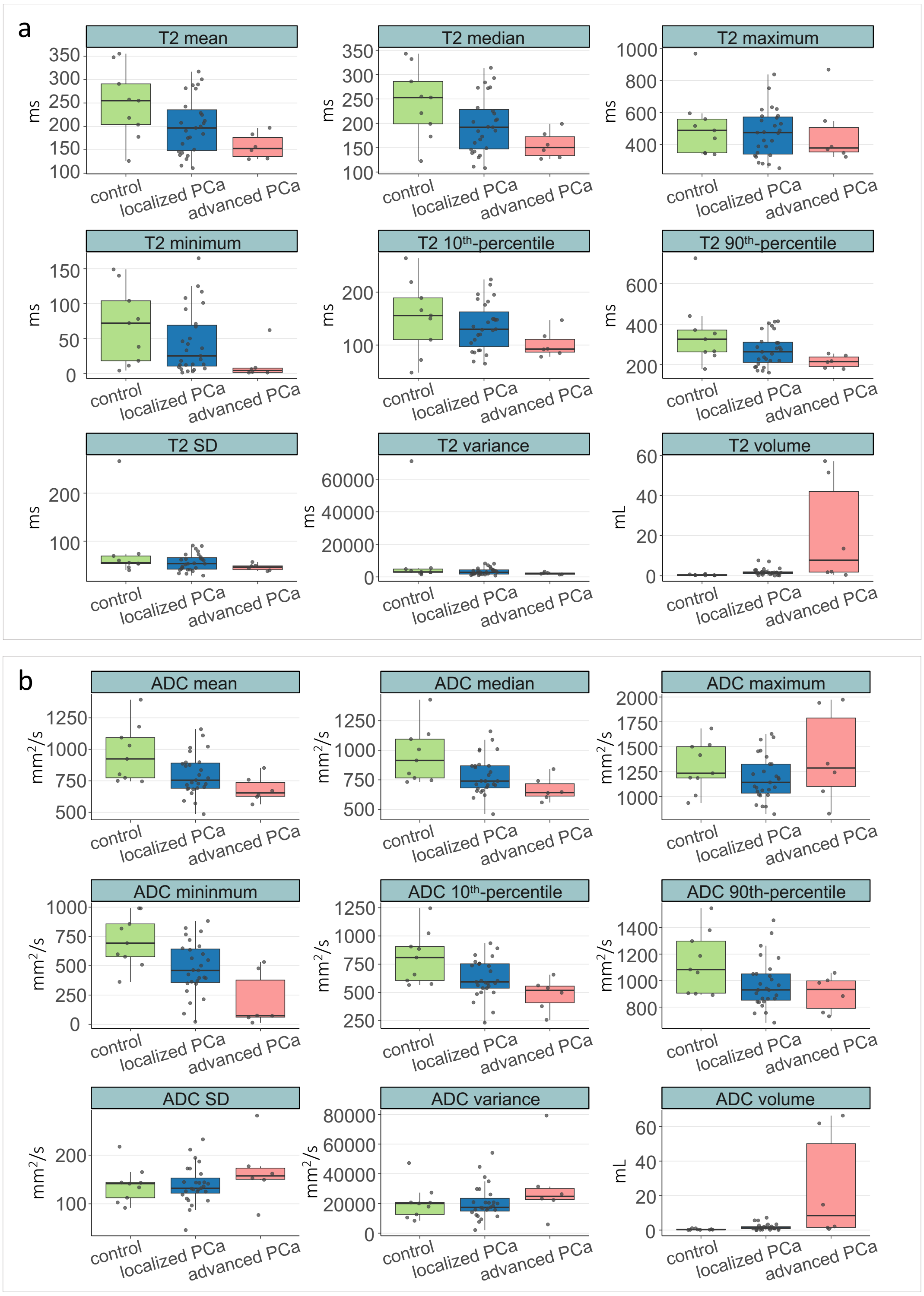


**Supplementary Figure S1:** Assessment of first-order radiological features from prostate MRI based on **a)** T2-weighted imaging or **b)** ADC mapping for 42 index lesions of 33 PCa patients and 9 cancer-free individuals. Comparison between cancer-free individuals and tumor patients, stratified for localized and advanced PCa. Box plot center lines indicate the median, and boxes illustrate the interquartile range with Tukey whiskers. Each dot represents one sample. Abbreviations: ADC = Apparent Diffusion Coefficient, MRI = magnetic resonance imaging, PCa = prostate cancer, SD = standard deviation

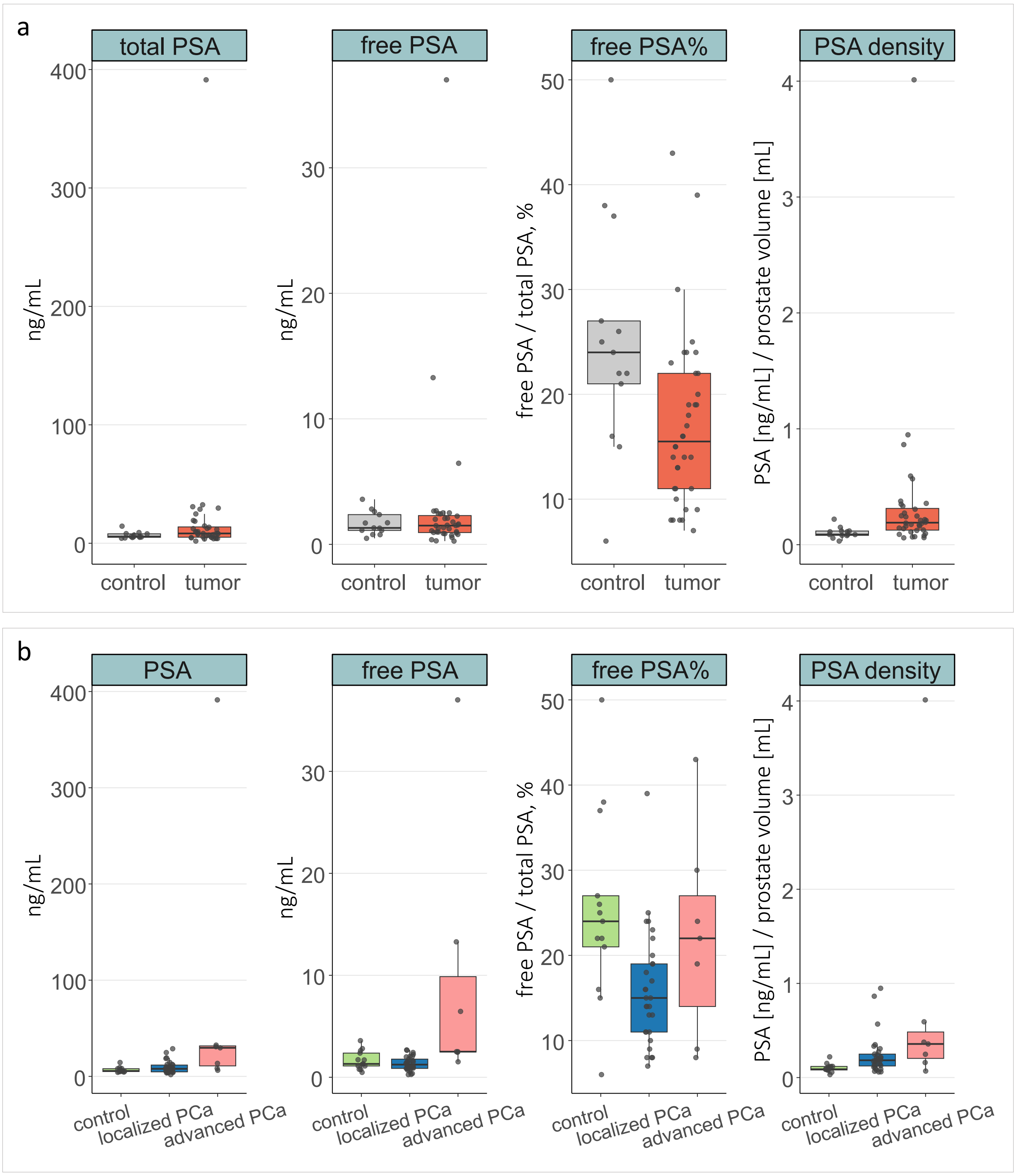


**Supplementary Figure S2:** Assessment of prostate-associated parameters measured in the blood from PCa patients and cancer-free individuals. **a)** Comparison between all PCa patients and cancer-free individuals. **b)** Comparison between cancer-free individuals and tumor patients, stratified for localized and advanced PCa. Box plot center lines indicate the median, and boxes illustrate the interquartile range with Tukey whiskers. Each dot represents one sample. Abbreviations: free PSA% = free-to-total PSA ratio, PSA = prostate-specific antigen

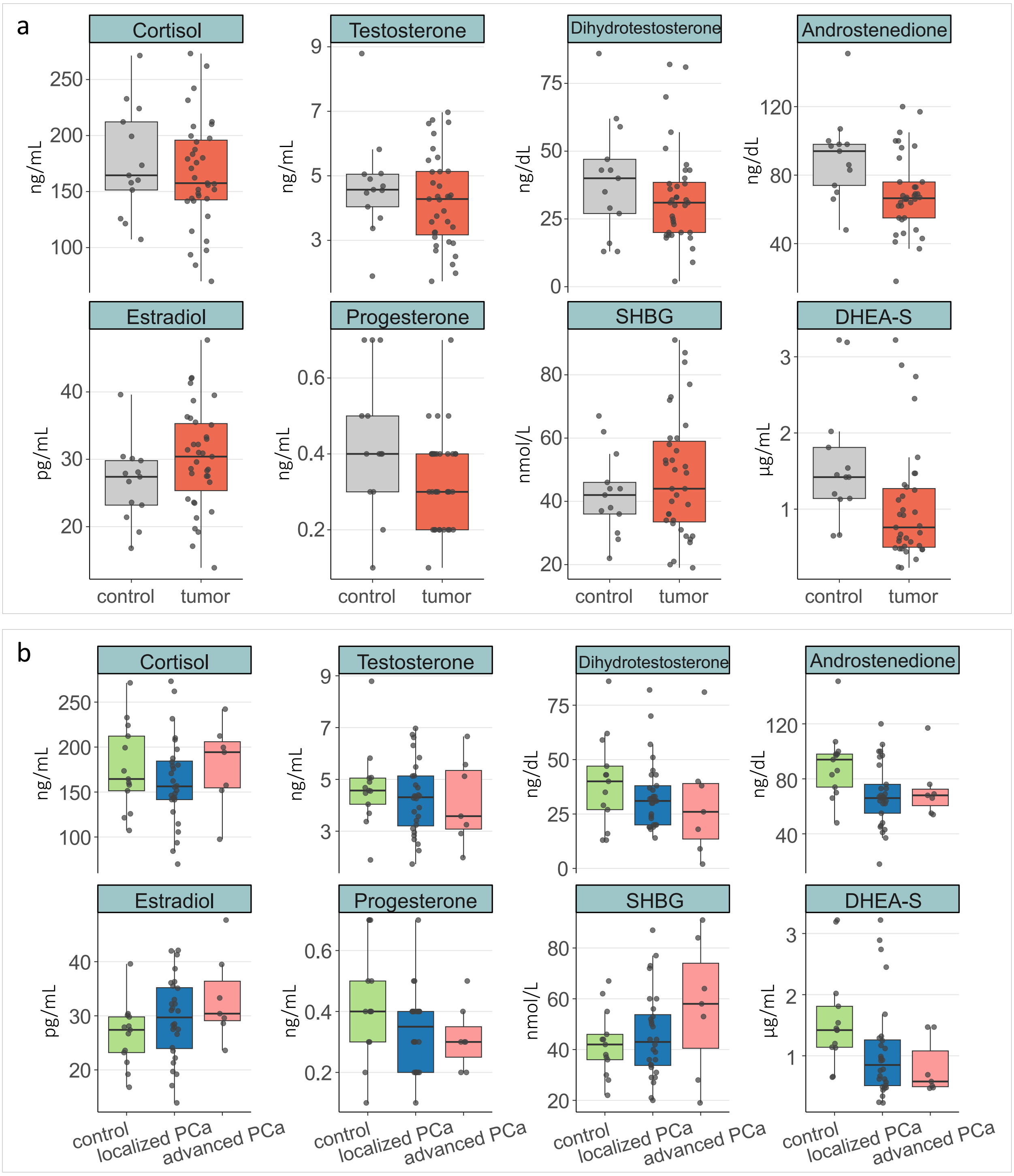


**Supplementary Figure S3:** Assessment of sex and adrenal hormones measured in the blood from PCa patients and cancer-free individuals. **a)** Comparison between all PCa patients and cancer-free individuals. **b)** Comparison between cancer-free individuals and tumor patients, stratified for localized and advanced PCa. Box plot center lines indicate the median, and boxes illustrate the interquartile range with Tukey whiskers. Each dot represents one sample. Abbreviations: DHEA-S = dehydroepiandrosterone sulfate, SHBG = sex hormone binding globulin

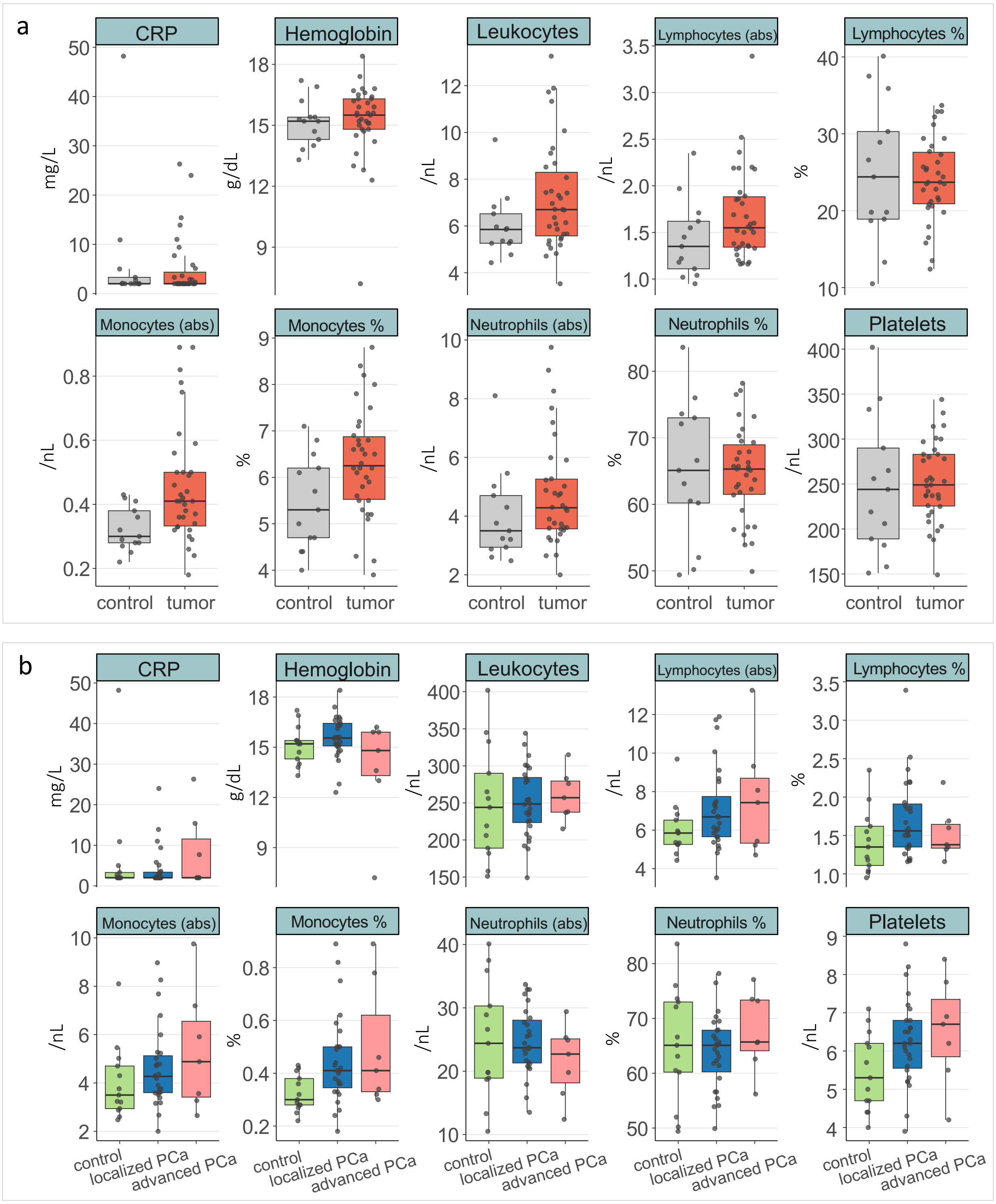


**Supplementary Figure S4:** Assessment of differential blood count and inflammation parameters measured in the blood from PCa patients and cancer-free individuals. **a)** Comparison between all PCa patients and cancer-free individuals. **b)** Comparison between cancer-free individuals and tumor patients, stratified for localized and advanced PCa. Box plot center lines indicate the median, and boxes illustrate the interquartile range with Tukey whiskers. Each dot represents one sample. Abbreviations: abs = absolute count, CRP = C-reactive protein

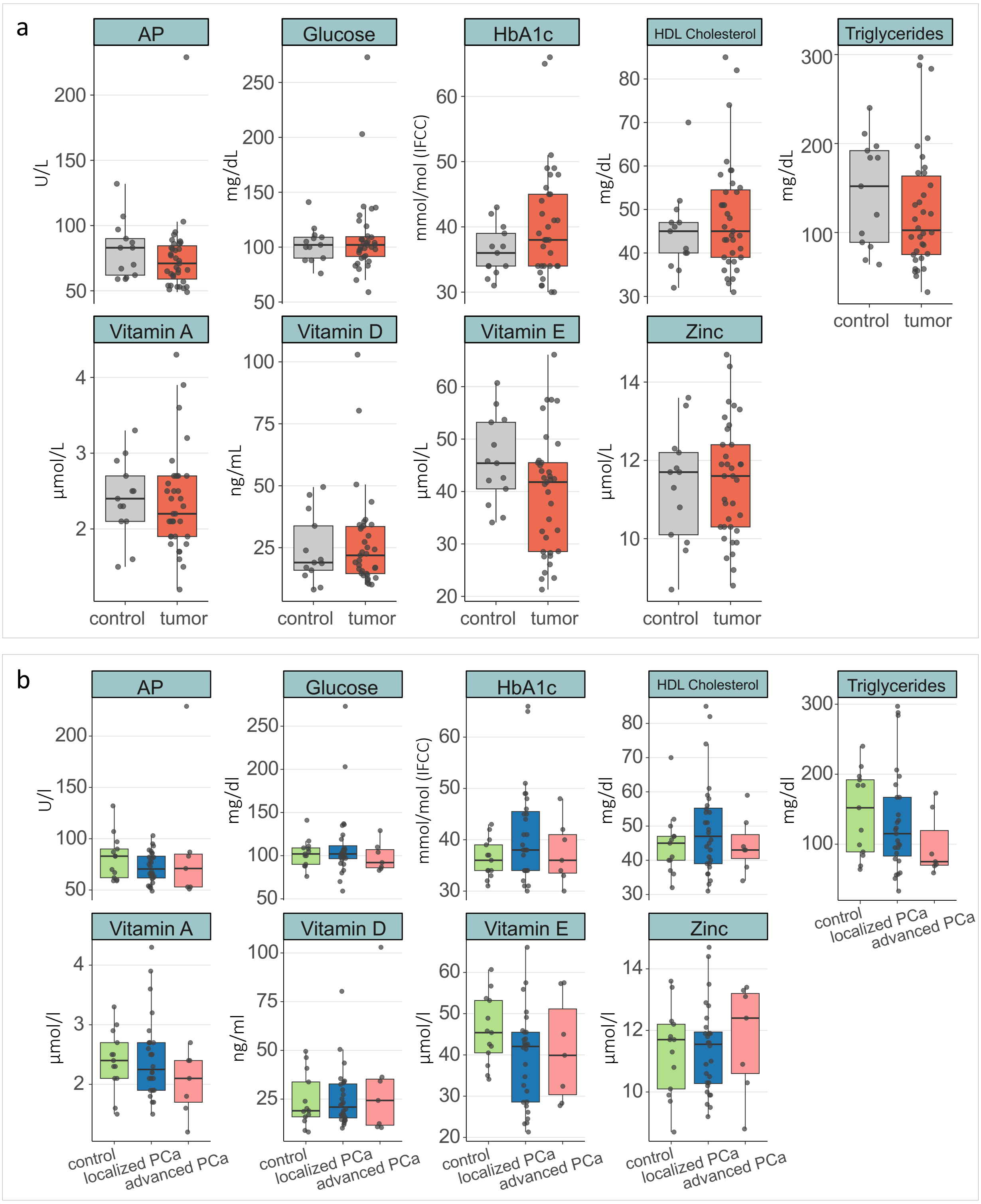


**Supplementary Figure S5:** Assessment of metabolic parameters and vitamins measured in the blood from PCa patients and cancer-free individuals. **a)** Comparison between all PCa patients and cancer-free individuals. **b)** Comparison between cancer-free individuals and tumor patients, stratified for localized and advanced PCa. Box plot center lines indicate the median, and boxes illustrate the interquartile range with Tukey whiskers. Each dot represents one sample. Abbreviations: AP = alkaline phosphatase, HbA1c = Hemoglobin A1c, HDL = high-density lipoprotein

a

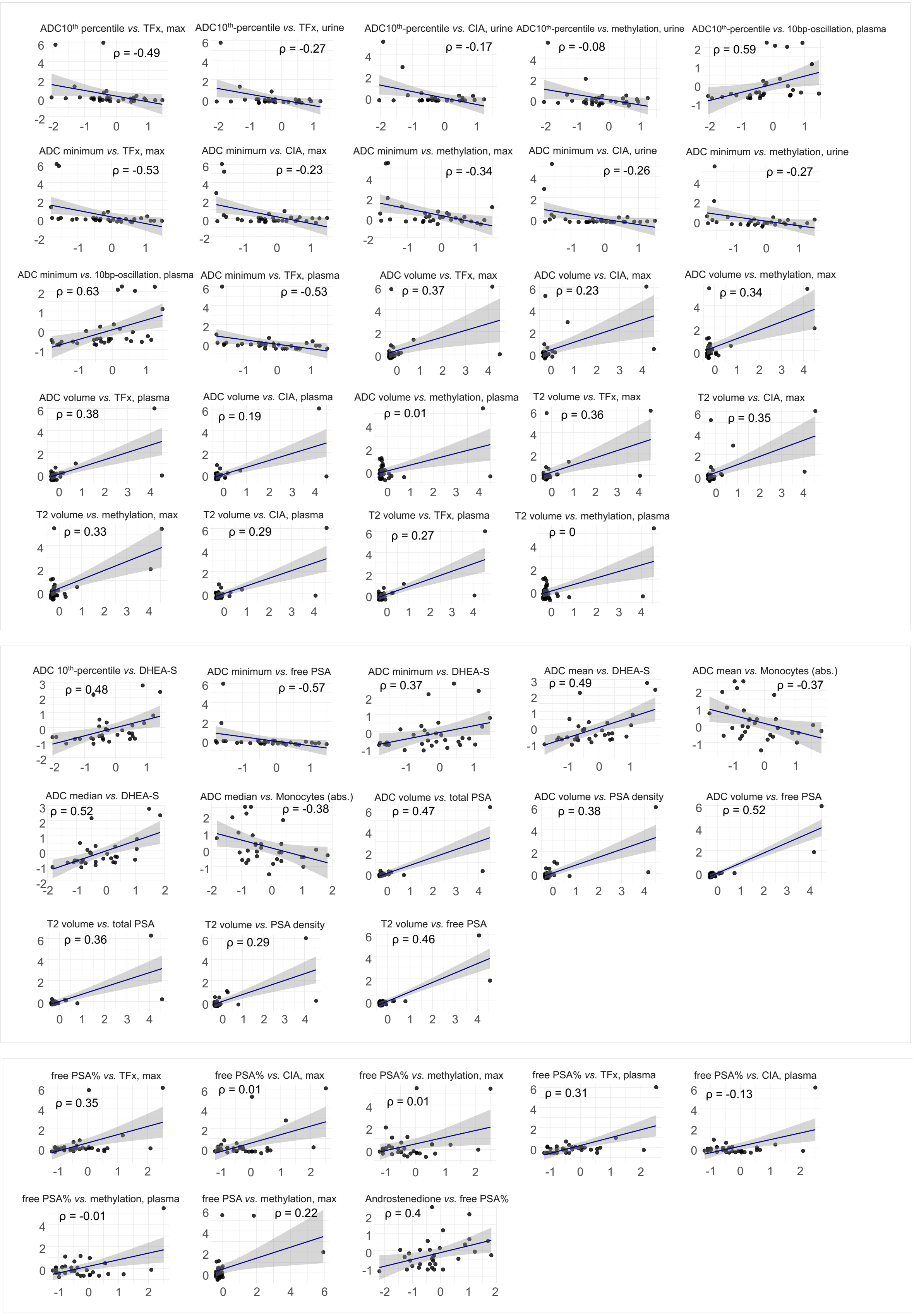

ADC10th percentile vs. TFx, max
ρ = -0.49
ADC10th-percentile vs. TFx, urine
ρ = -0.27
ADC10th-percentile vs. CIA, urine
ρ = -0.17
ADC10th-percentile vs. methylation, urine
ρ = -0.08
ADC10th-percentile vs. 10bp-oscillation, plasma
ρ = 0.59
ADC minimum vs. TFx, max
ρ = -0.53
ADC minimum vs. CIA, max
ρ = -0.23
ADC minimum vs. methylation, max
ρ = -0.34
ADC minimum vs. CIA, urine
ρ = -0.26
ADC minimum vs. methylation, urine
ρ = -0.27
ADC minimum vs. 10bp-oscillation, plasma
ρ = 0.63
ADC minimum vs. TFx, plasma
ρ = -0.53
ADC volume vs. TFx, max
ρ = 0.37
ADC volume vs. CIA, max
ρ = 0.23
ADC volume vs. methylation, max
ρ = 0.34
ADC volume vs. TFx, plasma
ρ = 0.38
ADC volume vs. CIA, plasma
ρ = 0.19
ADC volume vs. methylation, plasma
ρ = 0.01
T2 volume vs. TFx, max
ρ = 0.36
T2 volume vs. CIA, max
ρ = 0.35
T2 volume vs. methylation, max
ρ = 0.33
T2 volume vs. CIA, plasma
ρ = 0.29
T2 volume vs. TFx, plasma
ρ = 0.27
T2 volume vs. methylation, plasma
ρ = 0
ADC 10th-percentile vs. DHEA-S
ρ = 0.48
ADC minimum vs. free PSA
ρ = -0.57
ADC minimum vs. DHEA-S
ρ = 0.37
ADC mean vs. DHEA-S
ρ = 0.49
ADC mean vs. Monocytes (abs.)
ρ = -0.37
ADC median vs. DHEA-S
ρ = 0.52
ADC median vs. Monocytes (abs.)
ρ = -0.38
ADC volume vs. total PSA
ρ = 0.47
ADC volume vs. PSA density
ρ = 0.38
ADC volume vs. free PSA
ρ = 0.52
T2 volume vs. total PSA
ρ = 0.36
T2 volume vs. PSA density
ρ = 0.29
T2 volume vs. free PSA
ρ = 0.46
free PSA% vs. TFx, max
ρ = 0.35
free PSA% vs. CIA, max
ρ = 0.01
free PSA% vs. methylation, max
ρ = 0.01
free PSA% vs. TFx, plasma
ρ = 0.31
free PSA% vs. CIA, plasma
ρ = -0.13
free PSA% vs. methylation, plasma
ρ = -0.01
free PSA vs. methylation, max
ρ = 0.22
Androstenedione vs. free PSA%
ρ = 0.4

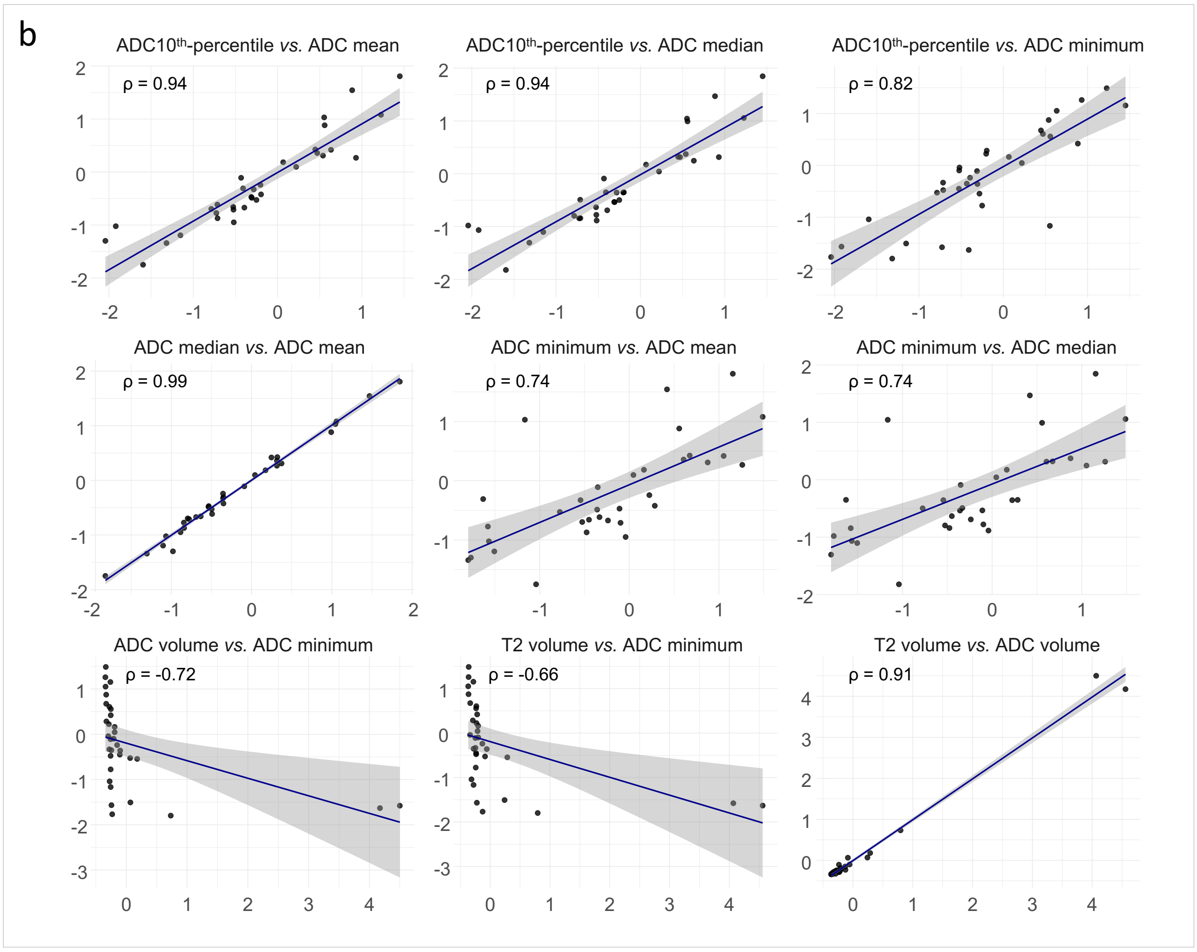
b
ADC10$^{th}$-percentile *vs.* ADC mean
ρ = 0.94
ADC10$^{th}$-percentile *vs.* ADC median
ρ = 0.94
ADC10$^{th}$-percentile *vs.* ADC minimum
ρ = 0.82
ADC median *vs.* ADC mean
ρ = 0.99
ADC minimum *vs.* ADC mean
ρ = 0.74
ADC minimum *vs.* ADC median
ρ = 0.74
ADC volume *vs.* ADC minimum
ρ = -0.72
T2 volume *vs.* ADC minimum
ρ = -0.66
T2 volume *vs.* ADC volume
ρ = 0.91

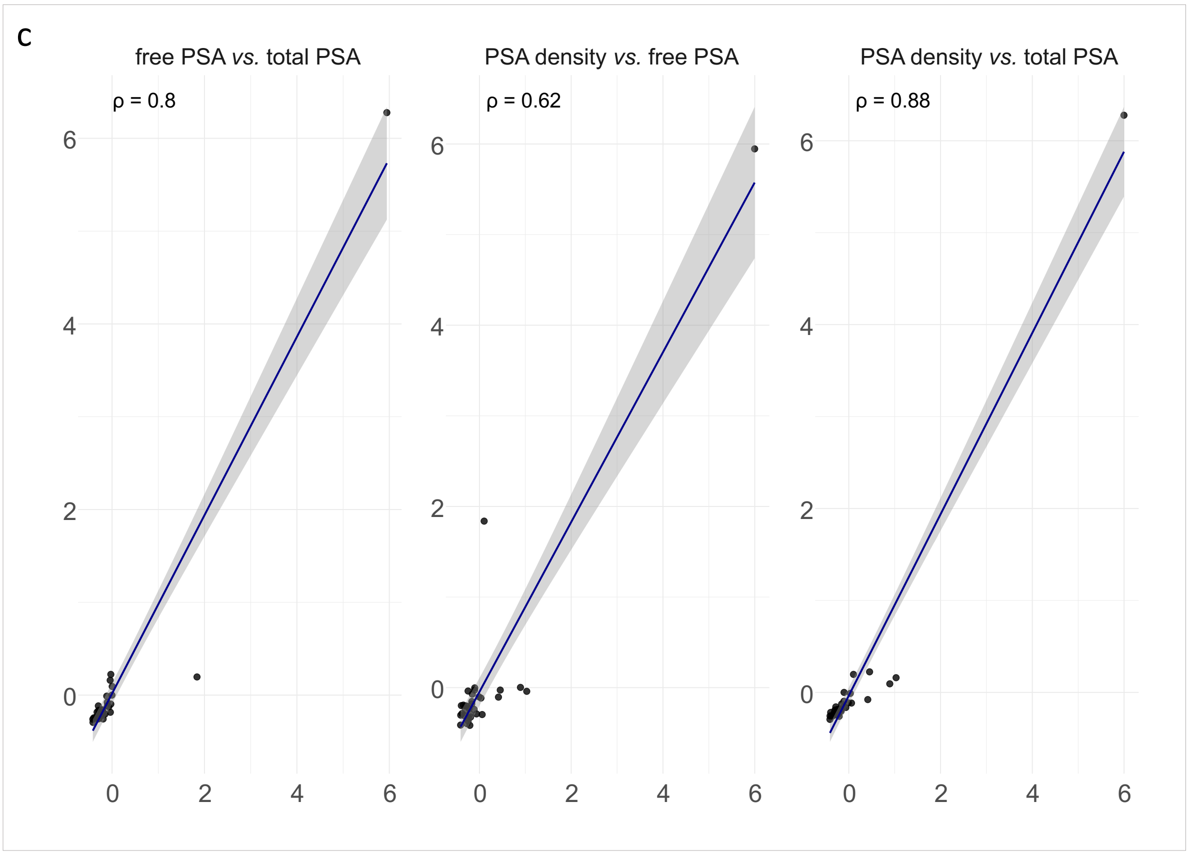
c
free PSA *vs.* total PSA
ρ = 0.8
PSA density *vs.* free PSA
ρ = 0.62
PSA density *vs.* total PSA
ρ = 0.88

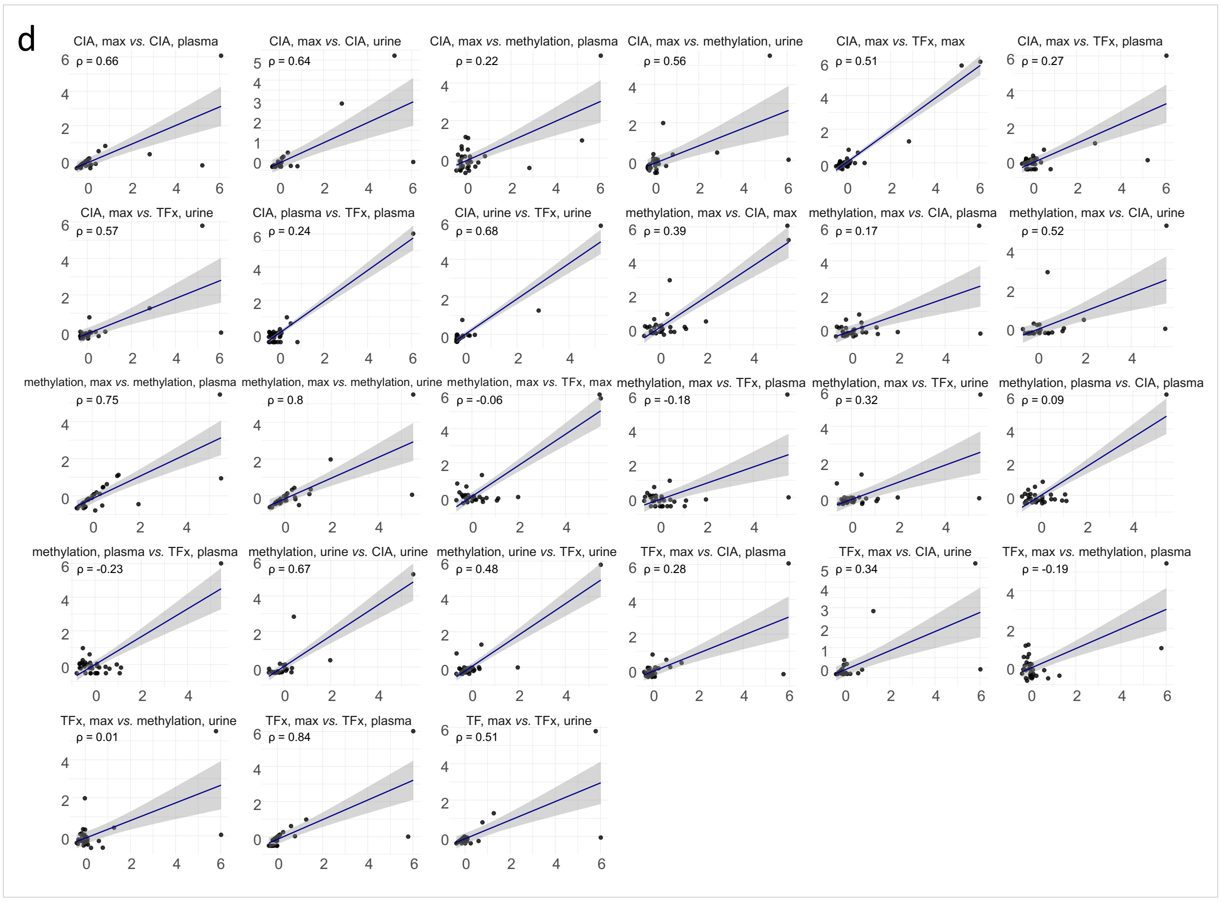


**Supplementary Figure S6:** Spearman correlation matrices of radiological features, serological parameters, and liquid biopsy markers in all PCa patients. Only patients with complete data sets across all modalities are included (n = 33). Each panel displays pairwise scatterplots of the indicated variables, with individual dots representing single patient samples. The black line denotes the fitted regression line from a Spearman rank correlation, and the grey shaded area represents the 95% confidence interval. Axes represent the raw measured values for each parameter. Four separate plots were generated: **a)** correlations across all three modalities, **b)** correlations between radiological features, **c)** correlations between serological parameters, and **d)** correlations between liquid biopsy markers. Abbreviations: CIA = chromosomal instability analysis, max = maximum, TFx = tumor fraction
